# Enhancing Cognitive Performance Prediction through White Matter Hyperintensity Connectivity Assessment: A Multicenter Lesion Network Mapping Analysis of 3,485 Memory Clinic Patients

**DOI:** 10.1101/2024.03.28.24305007

**Authors:** Marvin Petersen, Mirthe Coenen, Charles DeCarli, Alberto De Luca, Ewoud van der Lelij, Alzheimer’s Disease Neuroimaging Initiative, Frederik Barkhof, Thomas Benke, Christopher P. L. H. Chen, Peter Dal-Bianco, Anna Dewenter, Marco Duering, Christian Enzinger, Michael Ewers, Lieza G. Exalto, Evan F. Fletcher, Nicolai Franzmeier, Saima Hilal, Edith Hofer, Huiberdina L. Koek, Andrea B. Maier, Pauline M. Maillard, Cheryl R. McCreary, Janne M. Papma, Yolande A. L. Pijnenburg, Reinhold Schmidt, Eric E. Smith, Rebecca M. E. Steketee, Esther van den Berg, Wiesje M. van der Flier, Vikram Venkatraghavan, Narayanaswamy Venketasubramanian, Meike W. Vernooij, Frank J. Wolters, Xin Xu, Andreas Horn, Kaustubh R. Patil, Simon B. Eickhoff, Götz Thomalla, J. Matthijs Biesbroek, Geert Jan Biessels, Bastian Cheng

## Abstract

**Introduction:** White matter hyperintensities of presumed vascular origin (WMH) are associated with cognitive impairment and are a key imaging marker in evaluating cognitive health. However, WMH volume alone does not fully account for the extent of cognitive deficits and the mechanisms linking WMH to these deficits remain unclear. We propose that lesion network mapping (LNM), enables to infer if brain networks are connected to lesions, and could be a promising technique for enhancing our understanding of the role of WMH in cognitive disorders. Our study employed this approach to test the following hypotheses: (1) LNM-informed markers surpass WMH volumes in predicting cognitive performance, and (2) WMH contributing to cognitive impairment map to specific brain networks.

**Methods & results:** We analyzed cross-sectional data of 3,485 patients from 10 memory clinic cohorts within the Meta VCI Map Consortium, using harmonized test results in 4 cognitive domains and WMH segmentations. WMH segmentations were registered to a standard space and mapped onto existing normative structural and functional brain connectome data. We employed LNM to quantify WMH connectivity across 480 atlas-based gray and white matter regions of interest (ROI), resulting in ROI-level structural and functional LNM scores. The capacity of total and regional WMH volumes and LNM scores in predicting cognitive function was compared using ridge regression models in a nested cross-validation. LNM scores predicted performance in three cognitive domains (attention and executive function, information processing speed, and verbal memory) significantly better than WMH volumes. LNM scores did not improve prediction for language functions. ROI-level analysis revealed that higher LNM scores, representing greater disruptive effects of WMH on regional connectivity, in gray and white matter regions of the dorsal and ventral attention networks were associated with lower cognitive performance.

**Conclusion:** Measures of WMH-related brain network connectivity significantly improve the prediction of current cognitive performance in memory clinic patients compared to WMH volume as a traditional imaging marker of cerebrovascular disease. This highlights the crucial role of network effects, particularly in attention-related brain regions, improving our understanding of vascular contributions to cognitive impairment. Moving forward, refining WMH information with connectivity data could contribute to patient-tailored therapeutic interventions and facilitate the identification of subgroups at risk of cognitive disorders.

## Introduction

Cerebral small vessel disease (CSVD) is a major driver of vascular cognitive impairment (VCI) and often also contributes to dementia with a primary neurodegenerative or mixed pathology.^1^ White matter hyperintensities (WMH) are the signature imaging marker of CSVD, and mark sites of white matter disintegration caused by microangiopathic axonal loss and demyelination.^2,3^ However, a comprehensive understanding of mechanisms linking WMH to their broad range of clinical manifestations, specifically cognitive impairment, is still lacking.

Although there is a well-documented association between WMH volumes and cognitive functions at the group-level, the association between WMH volume and symptom severity demonstrates considerable variability with some individuals exhibiting fewer symptoms despite high WMH burden and vice versa.^4^ The apparent complexity of this relationship underscores the need for improved techniques for disease quantification to more accurately predict individual cognitive impairment for effective diagnostics and ultimately targeted treatment of CSVD patients.^5^ For example, lesion-symptom inference techniques have linked cognitive impairment to WMH located in strategic white matter regions, independent of total WMH volume.^4,6,7^

However, these recent findings might not fully reflect the complexity of CSVD-related cognitive impairment, which is thought to emerge from disturbances in the interplay of large-scale brain networks involving cortical and subcortical gray matter areas, interconnected by white matter tracts.^8^ In recent years, advanced imaging analysis models have been developed to comprehensively capture lesion effects on brain circuitry.^9^ Specifically, lesion network mapping (LNM) techniques capitalize on advanced neuroimaging to map lesions on reconstructions of the human brain network.^10^ By that, a lesion’s impact on connectivity to different brain regions can be quantified – i.e., the lesion’s network embedding is measured – allowing to infer which regions are disconnected. Application of LNM has been shown to predict clinical symptoms in a variety of neurological disorders that can be understood as “disconnection syndromes”, such as stroke or multiple sclerosis.^11,12^

Here, we propose LNM as a technique to quantify WMH-related, strategic neuronal disconnectivity for improved prediction of cognitive performance in CSVD. We employ LNM on a large-scale, multicenter dataset, integrating cognitive test results and MRI-based WMH segmentations from 3485 patients of 10 memory clinic cohorts through the Meta VCI Map Consortium.^6,13^ Our hypotheses are twofold: (1) LNM-based measures of WMH connectivity surpass WMH volumes in predicting cognitive performance, and (2) WMH contributing to cognitive deficits map to specific brain networks that functionally determine their symptom profile.

## Materials and methods

### Study population

Methodological details are illustrated in *figure 1*. We examined previously harmonized, cross-sectional clinical and imaging data of 3485 patients from 10 memory clinic cohorts of the Meta VCI Map Consortium.^6,13^ Meta VCI Map is a multi-site collaboration for conducting meta-analyses of strategic lesion topography in vascular cognitive impairment. The memory clinic cohorts included in this study comprise the Erasmus MC Memory Clinic Cohort (ACE, n=52, Netherlands), Alzheimer’s Disease Neuroimaging Initiative (ADNI, n=994, USA)^14^, UC Davis Alzheimer’s Disease Center Diversity Cohort (AUCD, n=641, USA)^15^, BrainIMPACT (n=53, Canada)^16^, Functional Assessment of Vascular Reactivity (FAVR, n=47, Canada)^16^, Harmonization (n=207, Singapore)^4^, Prospective Dementia Registry (PRODEM, n=367, Austria)^17^, TRACE-VCI (n=821, Netherlands)^18^, Utrecht Memory Clinic Cohort (UMCC, n=227, Netherlands) and VASCAMY (n=76, Germany). All cohorts include patients assessed at outpatient memory clinics for cognitive symptoms, undergoing structural MRI alongside neuropsychological tests of cognitive performance.

**Figure 1.**
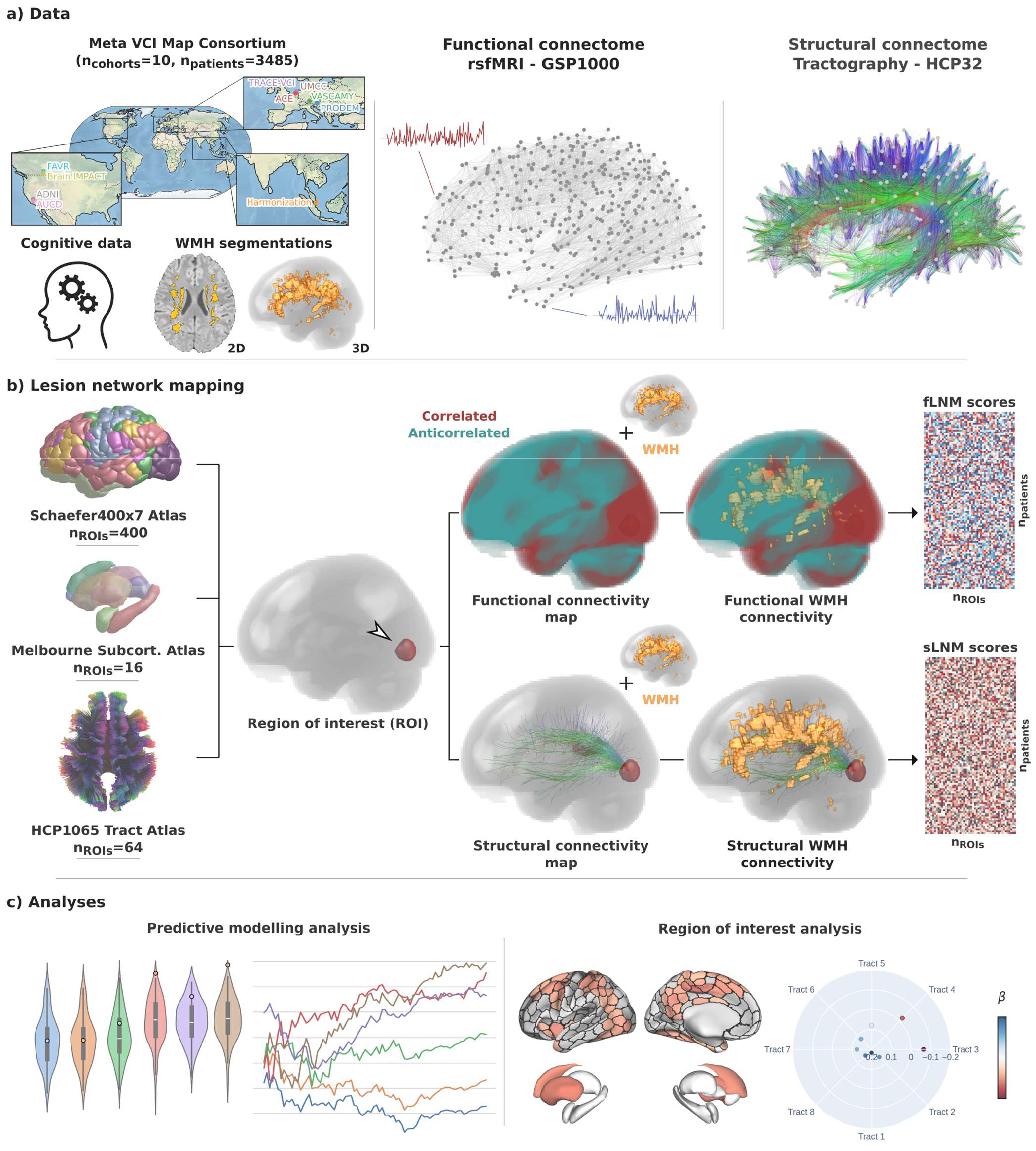
Methodology. a) Data from 10 memory clinic cohorts of the Meta VCI Map Consortium were used including harmonized cognitive scores and WMH segmentations in MNI space. For functional LNM we employed the GSP1000 normative functional connectome comprising resting-state fMRI data from 1000 healthy GSP participants. For structural lesion network mapping, we used the HCP32 normative structural connectome based on diffusion-weighted imaging data from 32 healthy HCP participants, detailing the fiber bundle architecture. b) LNM was performed to quantify the functional and structural connectivity of WMH to multiple ROIs (Schaefer400×7 cortical, Melbourne Subcortical Atlas subcortical, HCP1065 white matter areas). For this, voxel-level functional and structural connectivity maps were computed for each ROI, reflecting resting-state BOLD correlations or anatomical connection strength via tractography streamlines, respectively. ROIwise LNM scores were derived by averaging voxel-level connectivity indices within the normalized WMH masks, considering only positive correlation coefficients for functional mapping. This resulted in a matrix for both fLNM and sLNM scores per ROI per patient (n_ROIs_ x n_patients_). The matrices shown in the figure are populated with random data only serving as a visual aid. c) The fLNM and sLNM scores across patients were used in predictive models to estimate cognitive domain scores (predictive modelling analysis) and analyzed in permutation-based general linear models to identify regions significantly influencing the cognitive domain-WMH disconnectivity relationship at the ROI level (ROI-level inferential statistics). *Abbreviations*: fLNM = functional lesion network mapping, GSP = Genomic Superstruct Project, HCP = Human Connectome Project, ROI = region of interest, rsfMRI = resting-state functional magnetic resonance imaging, sLNM = structural lesion network mapping, WMH = white matter hyperintensities of presumed vascular origin.

Patients with cognitive impairment due to non-vascular, non-neurodegenerative causes (e.g., excessive alcohol use disorder, cerebral malignancies, multiple sclerosis) or monogenic disorders (e.g., CADASIL) were excluded. Further details on each cohort including sample-specific inclusion and exclusion criteria were reported previously.^6^

### Ethics approval

All cohorts received the requisite ethical and institutional approval in accordance with local regulations, which included informed consent, to allow data acquisition and sharing.^6^

### Cognitive assessments

Detailed harmonization procedures, including specific test-to-domain assignments, were reported previously.^19^ Neuropsychological tests from participating cohorts were norm-referenced against local norms or a healthy control group, and adjusted on the individual subject level for age, sex, and education. These tests were categorized into four cognitive domains: attention/executive function, information processing speed, language, and verbal memory. Within these domains, norm-referenced neuropsychological test scores were z-scored and averaged to obtain cognitive domain scores (z-scores), which capture individual-level cognitive domain performance relative to healthy controls.

### White matter hyperintensity segmentation

WMH segmentations were provided by the participating centers or performed at the UMC Utrecht (ACE cohort). Segmentation masks were obtained applying established automated neuroimaging software on fluid-attenuated inversion recovery (FLAIR) MRI.^20^ WMH segmentations were spatially normalized to the Montreal Neurological Institute (MNI)-152 template.^21^ To ensure registration quality, the normalized WMH masks were visually inspected and patients with failed registrations were excluded. Furthermore, random subsamples of normalized WMH segmentations were returned to the respective participating institutions to confirm the data quality. WMH segmentation masks were used to compute the total WMH volume as well as tract-level WMH volumes for each of the 64 white matter fiber tracts of the HCP1065 Tract Atlas.^22^ Details on cohort-specific segmentation and registration procedures were reported previously.^6,23^

### Lesion network mapping

LNM was performed to quantify the functional and structural connectivity of WMH to cortical, subcortical and white matter regions of interest (ROIs).^24^ ROIs were defined in MNI space according to the Schaefer400×7 Atlas (n_ROIs_=400), the Melbourne Subcortical Atlas (n_ROIs_=16) and the HCP1065 Tract Atlas (n_ROIs_=64) (*figure 1b*).^22,25,26^ For visualization of the investigated HCP1065 tracts, see *supplementary figure S1*.

Functional lesion network mapping (fLNM) was conducted using a normative functional connectome, derived from resting-state fMRI scans of 1,000 healthy individuals from the Genomic Superstruct Project (GSP1000).^27^ Preprocessing included global signal regression and spatial smoothing at a 6mm full width at half maximum kernel as previously detailed.^28^ For each ROI, we averaged blood oxygen level-dependent (BOLD) signal fluctuations across voxels within the ROI and correlated this aggregate time series with BOLD signals of all brain voxels. This process generated 1,000 Pearson correlation coefficients per voxel, i.e., one for each GSP1000 subject, which were then Fischer z-transformed and averaged across subjects to create a functional connectivity map per ROI. Functional connectivity map computations were performed using the ROI masks as seeds in the *connectome mapper* function of Lead-DBS (<u>lead-dbs.org</u>).^29^ Subsequently, ROI-level fLNM scores were calculated by averaging positive Pearson correlation coefficients within the WMH mask, reflecting each ROI’s functional connectivity to WMH.

Structural lesion network mapping (sLNM) was performed employing a normative structural connectome of 32 subjects of the Human Connectome Project (HCP32).^30^ The structural connectome was reconstructed by applying DSI Studio on multi-shell diffusion MRI data acquired on a MRI scanner specifically designed for high-fidelity connectome reconstruction. Streamlines resulting from whole brain tractography were normalized to MNI and aggregated across subjects.^31^ Employing Lead-DBS, voxel-wise structural connectivity maps were computed per atlas ROI, quantifying per voxel the number of streamlines connecting the voxel to the ROI.^29^ ROI-level sLNM scores, reflecting structural connectivity between WMH and individual ROI, were determined by averaging the voxel values (representing streamline counts to the ROI) within the WMH mask.

Summarized, LNM yielded both a fLNM and sLNM score for each ROI per subject, indicating the functional and structural connectivity between WMH and ROI, respectively.

### Predictive modelling analysis

To evaluate the predictive capacity of fLNM and sLNM scores, we performed a predictive modelling analysis leveraging scikit-learn (v. 1.0.2, <u>scikit-learn.org</u>) and julearn (v. 0.3.0, <u>juaml.github.io/julearn</u>).^32,33^ This work defines ‘prediction’ in accordance with previous studies as the estimation of target variables using a trained statistical model on new unseen data – emphasizing the crucial aspect of model generalizability.^9,34,35^ We note that this definition varies from those indicating longitudinal study designs used in epidemiological contexts.^36^ In the analysis, six different feature sets were compared: (1) demographics (age, sex and education), (2) total WMH volume + demographics, (3) tract-level WMH volumes + demographics, (4) ROI-level fLNM scores + demographics, (5) ROI-level sLNM scores + demographics, (6) ROI-level fLNM and sLNM scores + demographics.

For each cognitive domain, multivariable ridge regression models were trained using the abovementioned feature sets to predict cognitive domain scores. Ridge regression models include a L2 penalty that reduces coefficients to mitigate overfitting and address multicollinearity. We optimized the L2 penalties through a 10-fold nested cross-validation, tuning α values ranging from 0.001 to 1000 (α = 0.001, 0.01, 0.1, 1, 10, 100, 1000). The model performance was scored by the Pearson correlation between actual and predicted cognitive domain scores, supplemented with explained variance (R^2^, coefficient of determination) and negative mean squared error as additional measures of performance. In line with best practices, explained variance was calculated via sum-of-squares formulation (using scikit-learn’s *r2_score*) instead of squaring Pearson correlations.^34^ Before model fitting, continuous input features were z-scored in a cross-validation consistent manner to avoid data leakage.^37^ To maintain a consistent distribution of the target variable across training and test sets, we employed julearn’s *ContinuousStratifiedKFold* function for creating the folds. Cross-validations were repeated 10 times with varied random splits to minimize bias from any single split.^38^ This approach yielded 100 scores for each feature-target set combination which were compared between feature sets using a machine learning-adjusted t-test.^39^ We repeated the predictive modelling analysis for different sample sizes (20%-100%, 1% steps, randomly sampled) to examine the robustness and sample size dependency of predictive performances. As a whole, this analysis follows current best practices of predictive modelling in neuroimaging.^34^

### Region of interest-level inferential statistics

To investigate whether WMH-related connectivity of specific brain circuits links to impaired cognitive performance, we conducted permutation-based testing for linear associations between regional LNM scores and cognitive domain scores in a general linear model. All statistical analyses were conducted in FSL’s Permutation Analysis of Linear Models (PALM) based on MATLAB (v. 2021b) and Python 3.9.1 leveraging neuromaps (v. 0.0.5).^40–42^ Statistical tests were two-sided (n_permutation_=5000), with a p<0.05 as the significance threshold. To account for multiple comparisons, p*-* values were adjusted for family-wise error. General linear models were adjusted for age, sex and education. To obtain standardized *β*-coefficients, input variables were z-scored beforehand. As a result, *β*-coefficients and p-values were obtained for each cortical, subcortical, and white matter ROI (n_ROIs_=480) indicating the strength and significance of the LNM score’s linear association with cognitive domain scores for each ROI. To aid in interpreting the spatial effect patterns, we averaged the *β*-coefficients representing cortical effects in the 7 intrinsic resting-state networks (Yeo networks), which reflect the cerebral cortex’s intrinsic functional organization.^28^ The Schaefer400×7 Atlas assigns ROIs to these networks: visual, somatomotor, dorsal attention, ventral attention (salience), limbic, frontoparietal control, and default mode network.^25^ Significance was tested via spin permutations (n_spins_=1000) which represent a null model preserving the inherent spatial autocorrelation of cortical information.

### Sensitivity analyses

During computations of fLNM scores, we decided to only consider positive Pearson correlations of resting-state BOLD signal within WMH masks following previous approaches as the role of negative correlations is controversial.^43^ However, some studies suggest biological meaning in anticorrelations of BOLD signal fluctuations.^44,45^ Hence, we conducted a sensitivity analysis based on fLNM scores computed by averaging only the negative Pearson correlations in the WMH masks. We reconducted the predictive modelling analysis and ROI-level inferential statistics using these negative fLNM scores.

Moreover, previous work employs thresholding to discard potentially noisy connectivity information. To further examine the effect of thresholding on our results we repeated the predictive modelling analysis comparing the main analysis results to fLNM and sLNM scores computed based on 25% and 50% highest voxel intensities in the WMH mask. For negative fLNM scores, the lowest 25% and 50% voxel intensities in the WMH mask were considered.

### Exploratory analyses

Further exploratory analyses including investigations of voxel-level lesion network maps and structure-function coupling of LNM scores are described in *supplementary text S2*.

### Data availability

Analysis code can be accessed on GitHub (https://github.com/csi-hamburg/2024_petersen_wmh_disconnectivity_memory_clinic). The data that support the findings of this study are available from the corresponding author/project leads on reasonable request (https://metavcimap.org/group/become-a-member/). Restrictions related to privacy and personal data sharing regulations and informed consent may apply.

## Results

### Sample characteristics

The pooled study sample of 3485 patients had a mean age of 71.7 ± 8.9 years and 49.8% were female. Among patients, 777 (22.3%) had subjective cognitive impairment, 1389 (39.9%) had mild cognitive impairment, and 1319 (37.9%) had dementia. Further details on the sample characteristics can be found in *table 1*. A heatmap of WMH distribution can be found in *supplementary figure S3*.

**Table 1.** Sample characteristics.

| Metric | Stat |
| --- | --- |
| Age in years, mean ± SD (n) | 71.71 ± 8.87 (3485) |
| Female, n (%) | 1737 (49.8) |
| Years of education, mean ± SD (n) | 12.89 ± 4.45 (3485) |
| **Patient ethnicity** |  |
| Afrocaribbean, n (%) | 198 (5.7) |
| Asian, n (%) | 237 (6.8) |
| Caucasian / European / White, n (%) | 1620 (46.5) |
| Hispanic, n (%) | 146 (4.2) |
| Other, n (%) | 52 (1.5) |
| **Diagnosis** |  |
| Subjective cognitive impairment | 777 (22.30) |
| Mild cognitive impairment | 1389 (39.86) |
| Dementia | 1319 (37.85) |
| **For dementia cases: probable etiology** |  |
| Alzheimer's dementia, n (%) | 764 (57.9) |
| Vascular dementia, n (%) | 85 (6.4) |
| Frontotemporal dementia, n (%) | 44 (3.3) |
| Dementia with Lewy bodies, n (%) | 24 (1.8) |
| **Cardiovascular risk factors** |  |
| Current smoking, n (%) | 499 (14.3) |
| Previous smoking, n (%) | 459 (13.2) |
| Hypertension, n (%) | 1714 (49.2) |
| Hypercholesterolemia, n (%) | 1098 (31.5) |
| Diabetes mellitus, n (%) | 492 (14.1) |
| BMI, mean ± SD (n) | 25.28 ± 4.75 (640) |
| **Comorbidities** |  |
| Atrial fibrillation, n (%) | 98 (2.8) |
| History of prior stroke, n (%) | 244 (7.0) |
| History of prior transient ischemic attack (TIA), n (%) | 62 (1.8) |
| History of prior other vascular events, n (%) | 715 (20.5) |
| **Imaging** |  |
| WMH volume in ml, median [IQR] (n) | 6.19 [14.21] (3485) |
| **Cognitive function** |  |
| Mini-mental state examination, mean ± SD (n) | 25.0 ± 4.7 (3327) |
| Attention / executive function, z, mean ± SD (n) | -1.12 ± 1.10 (3446) |
| Information processing speed, z, mean ± SD (n) | -0.96 ± 1.61 (2417) |
| Language, z, mean ± SD (n) | -1.08 ± 1.86 (2041) |
| Verbal memory, z, mean ± SD (n) | -1.48 ± 1.30 (3242) |
| Abbreviations: ml = milliliter, SD – standard deviation, z – harmonized z-score |  |

### Predictive modelling analysis

To evaluate if information on WMH network connectivity exceeds the predictive capacity of volumetric WMH metrics for cognitive performance, we first computed regional fLNM and sLNM scores, that capture the structural and functional connectivity profile of WMH. We then employed ridge regression for predictive modelling. Model performance was assessed via Pearson correlation (r) of predicted and actual cognitive domain scores averaged across folds. All models incorporated age, sex, and education (demographics) as features to establish a performance baseline. The corresponding results are visualized in *figure 2a*. In summary, LNM scores significantly improved cognitive function prediction in all domains, except language, over WMH volumes. In detail, the predictive performance achieved by the demographics-only model was r = 0.312 for attention / executive function, r = 0.239 for information processing speed, r = 0.404 for language, and r = 0.305 for verbal memory. Models informed by total or tract-wise WMH volumes achieved a predictive performance of r = 0.341 - 0.365 for attention / executive function, r = 0.247 – 0.250 for information processing speed, r = 0.404 – 0.416 for language, and r = 0.327 – 0.356 for verbal memory. For the prediction of attention / executive function, models informed by LNM scores exhibited a significantly higher predictive performance than models informed by volumetric WMH measures (LNM: r = 0.399 - 0.410 vs. WMH volume: r = 0.341 – 0.365; adjusted t-test, all p < 0.05). LNM-informed models also better predicted information processing speed (LNM: r = 0.310 - 0.316 vs. WMH volume: r = 0.247 – 0.250, adjusted t-test, all p < 0.05) as well as verbal memory (LNM: r = 0.390 - 0.408 vs. WMH volume: r = 0.327 – 0.356; adjusted t-test, all p < 0.05). Across these domains, the best prediction was achieved by models incorporating both structural and functional LNM scores. For attention / executive function, comparing the improvement from the demographics-based model to the model informed by total WMH volume (0.341 – 0.312 = 0.029) with the improvement to the model based on both LNM modalities (0.410 – 0.312 = 0.098), the usage of fLNM and sLNM scores amounts to a 3.38-fold increase (0.098 / 0.029 = 3.38) in added predictive performance. Considering both LNM modalities for predicting information processing speed and verbal memory amounted to 7.00-fold and 4.68-fold increase in predictive performance, respectively. For the prediction of language domain scores, performance between LNM-informed models and WMH volume measures did not differ significantly (LNM: r = 0.380 - 0.409 vs. WMH volume: r = 0.404 – 0.416, all p > 0.05). See *supplementary materials S4* and *S5* for predictive modelling results using explained variance and negative mean squared error as scoring methods. Details on regional averages of LNM scores are shown *supplementary figure S6*.

**Figure 2.**
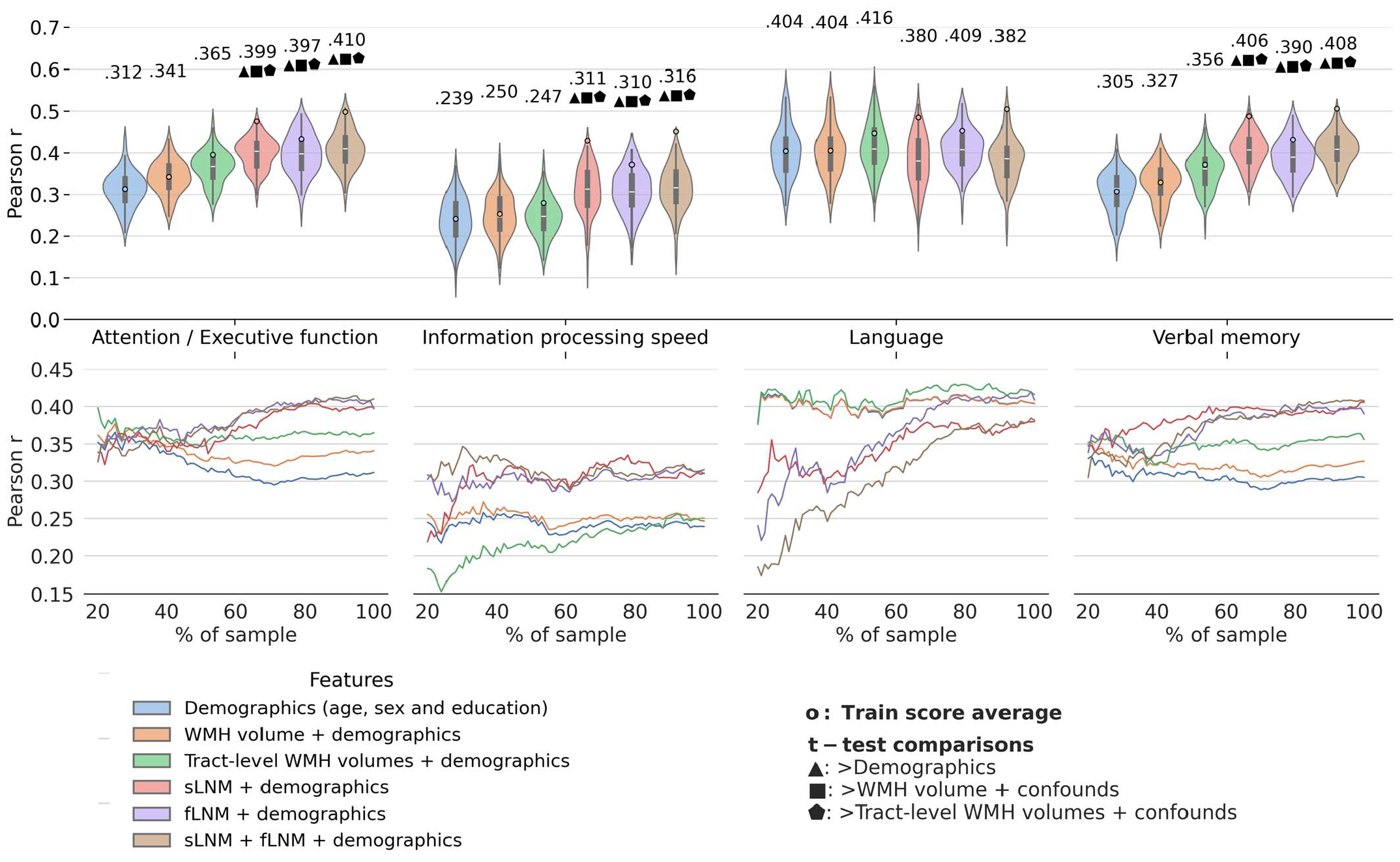
Predictive modelling analysis. Violin plots illustrate prediction outcomes across cognitive domains. Each violin displays the distribution of Pearson correlations (between actual and predicted cognitive domain performance; 10-fold cross-validation × 10 repeats = 100 folds → 100 Pearson correlations) for a model informed by a different feature set. The higher the Pearson correlation, the higher the prediction performance. blue: de-mographics (age, sex and education); orange: total WMH volume + demographics; green: tract-level WMH volumes + demographics; red: sLNM scores + demographics; purple: fLNM scores + demographics; brown: sLNM scores + fLNM scores + demographics. Average Pearson correlations are indicated above each violin, with colored dots showing training score averages. Geometric symbols denote t-test results comparing LNM-based models against demographics- and WMH volume-based models: ▴ indicates higher Pearson correlation than demographics, ▪ than WMH volume + demographics, ☗ than tract-level WMH volume + demographics. Below the violin plots, performance curves display the average Pearson correlations across folds, for subsets randomly sampled in sizes ranging from 20% to 100% of the entire dataset. Line colors match the corresponding violin plots in panel a) which display predictive modelling results for the full sample size. Again, higher Pearson correlation indicates higher prediction performance. *Abbreviations*: fLNM = functional lesion network mapping, sLNM = structural lesion network mapping, WMH = white matter hyperintensities of presumed vascular origin.

To test the robustness of prediction results, we repeated the analysis in randomly chosen subsamples of increasing sizes (*figure 2b*). For attention / executive function and verbal memory, LNM-informed models started to consistently exceed WMH volume-based models at approximately 50% (attention / executive function: n=1723, verbal memory: n=1712; note that data availability differed between cognitive domain scores) of the sample size.

For information processing speed, LNM-informed models surpassed WMH volume-based models at approximately 25% (n=604) of the sample size. Regarding language, LNM-informed models approximated the performance of WMH volume-based models with increasing sample sizes. For all cognitive domain scores, predictive performance in the sample size range 80-100% showed high stability and only minor increases indicating saturation.

### Contextualization of WMH connectivity: Region of interest analysis

We tested if WMH connectivity of specific brain circuits links to cognitive performance by quantifying the association between *regional* LNM scores (cortical brain regions and white matter tracts) and cognitive domain scores adjusting for age, sex and education.

Results of the general linear model linking LNM scores in cortical and subcortical gray matter regions to cognitive domain scores are shown in *figure 3*. Higher fLNM scores (i. e. increased WMH connectivity) in cortical regions of the dorsal attention and ventral attention networks were linked to lower attention / executive function and verbal memory (*figure 3a* and *c*). Regarding information processing speed, the extent of the effect was limited to several cortical brain areas mapping to the dorsal attention network (*figure 3b*). In terms of sLNM, higher scores in the dorsal attention network were significantly associated with lower attention / executive function and information processing speed (*figure 3d* and *e*). Again, information processing speed showed a spatially more limited effect pattern. The relationship of regional sLNM and verbal memory scores showed a different spatial distribution mapping to the ventral attention, frontoparietal and default mode network (*figure 3f*). The cortical and subcortical LNM scores showed no significant association with the language domain score.

**Figure 3.**
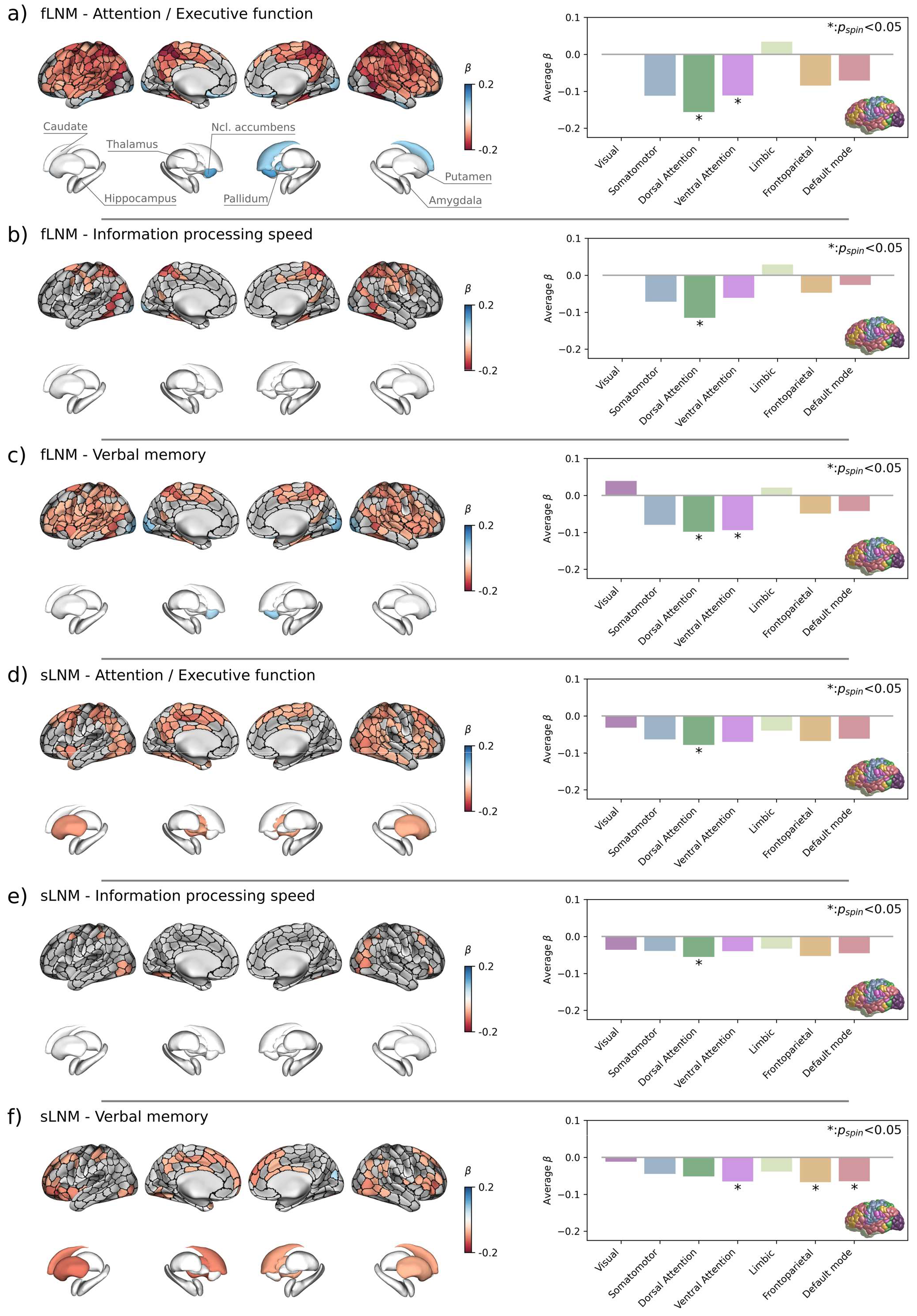
Inferential statistics results of cortical and subcortical gray matter. Anatomical plots on the left side display the regional relationship between LNM scores and cognitive domain scores. ROIs in which LNM scores across participants were significantly associated with cognitive domain scores after family-wise error-correction are highlighted by colors encoding *β*-coefficients from general linear models: a negative *β* (red) denotes that a higher regional LNM score, i.e., higher WMH connectivity, is associated to a lower performance in individual cognitive domains; a positive *β* (blue) indicates that a higher regional LNM score is linked to a higher cognitive domain performance. Barplots on the right side display the corresponding *β* coefficients averaged in the canonical (Yeo) resting-state functional connectivity networks. The brain in the lower right corner indicates the regional distribution of the canonical resting-state networks with colors corresponding to the bars. Statistical significance was assessed using spin permutations. Each row corresponds with a different combination of lesion network mapping modality and cognitive domain: a) fLNM – attention / executive function, b) fLNM – information processing speed, c) fLNM – verbal memory, d) sLNM – attention / executive function, e) sLNM – information processing speed, f) sLNM – verbal memory. *Abbreviations*: fLNM = functional lesion network mapping, *p*_*spin*_ = p-value derived from spin permutations, ROIs = regions of interest, sLNM = structural lesion network mapping.

The results for anatomically predefined white matter tracts are shown in *figure* 4. For tract-level fLNM, lower cognitive performance in attention / executive function, information processing speed and verbal memory was most strongly linked to higher fLNM scores in association and projection tracts connecting the parietal cortex (*figure 4b*): the middle longitudinal fasciculus (MdLF), parietal corticopontine tract (CPT), dorsal, medial and ventral sections of the superior longitudinal fasciculus (SLF 1-3), the parieto-parahippocampal cingulate (C parietoparahipp.). For attention / executive function, a strong negative effect was also evident for the right arcuate fasciculus (AF). For verbal memory, significant negative effects were additionally found for the corticobulbar tract (CBT) and frontal aslant tract (FAT).

**Figure 4.**
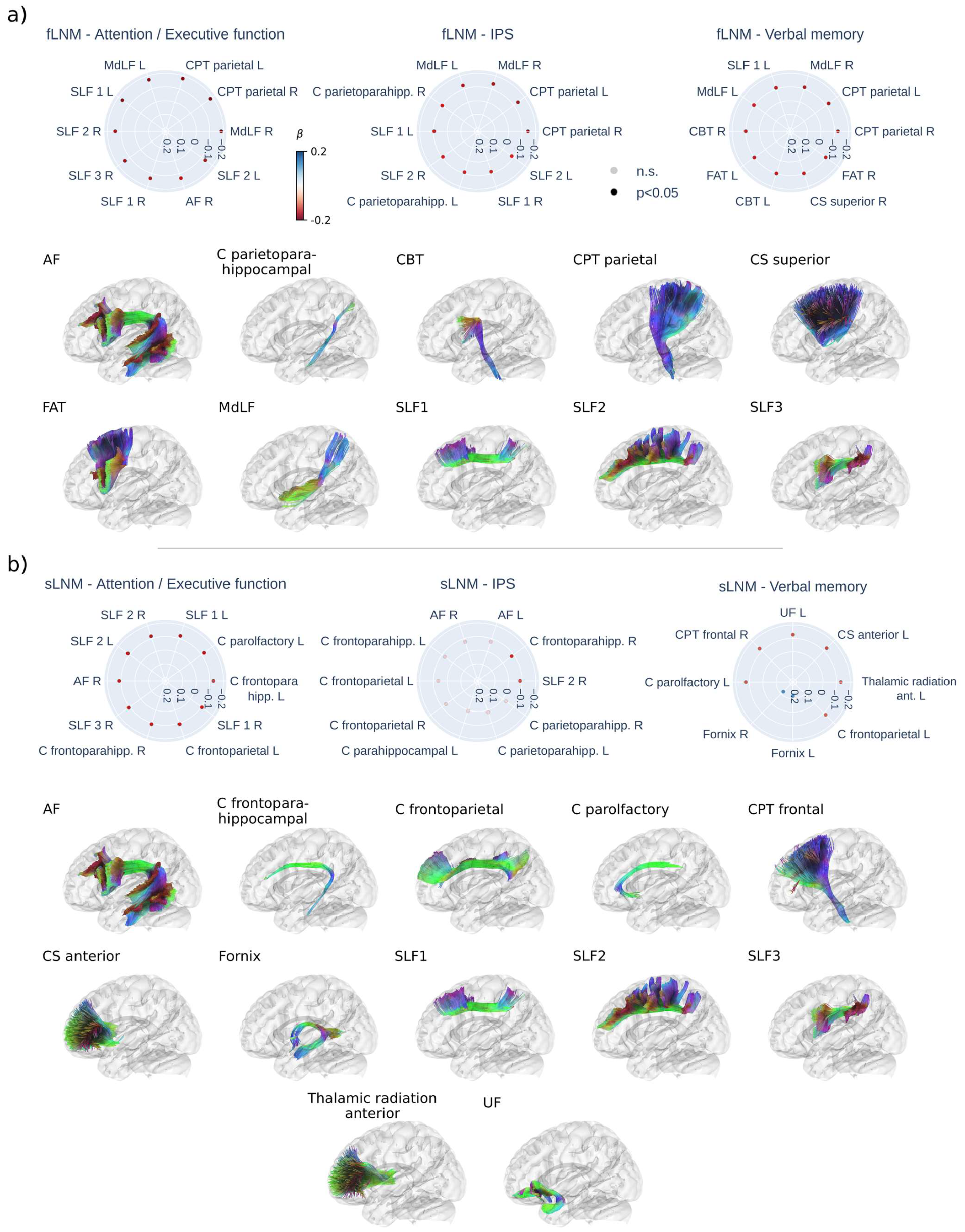
Inferential statistics results of white matter tracts. Radar plots displaying the top 10 of strongest linear associations (standardized *β*) for the functional (a) and structural (b) lesion network mapping scores in each tract in association with cognitive domain scores. Strongest associations are shown at the 3 o’clock position, decreasing in strength counterclockwise. Red dots indicate a negative association (higher LNM score – lower cognitive domain score) and blue dots indicate a positive association (higher LNM score – higher cognitive domain score). Faintly colored dots indicate non-significant associations. Tracts with a significant association are displayed below the radar plots in alphabetical order. For paired tracts only left side examples are visualized. *Tract abbreviations*: AF = arcuate fascicle, C = cingulate, CBT = corticobulbar tract, CPT = corticopontine tract, CS = corticostriatal pathway, F = fornix, FAT = frontal aslant tract, MdLF = middle longitudinal fasciculus, SLF = superior longitudinal fasciculus, UF = uncinate fasciculus; *Abbreviations*: fLNM = functional lesion network mapping, IPS – information processing speed, n.s. = non-significant, p = p-value, sLNM = structural lesion network mapping.

Regarding tract-level sLNM, lower attention / executive function and verbal memory were significantly associated with higher sLNM scores in association and projection tracts connecting frontal regions (*figure 4c*): the frontoparahippocampal cingulate (C parietoparahipp.), parolfactory cingulate (C parolfactory), the superior longitudinal fasciculus (SLF 1-3), frontoparietal cingulate (C frontoparietal), anterior thalamic radiation, anterior corticostriatal pathways (CS anterior), uncinate fascicle, frontal corticopontine tract (CPT frontal). For attention / executive function, a strong negative effect was also evident for the right arcuate fasciculus (AF). Furthermore, higher verbal memory scores were significantly linked to higher sLNM scores in the fornices. Information processing speed showed a significant negative association with sLNM scores in the right medial superior longitudinal fasciculus (SLF 2) and frontoparahippocampal cingulate (C frontoparahipp.). Tract-level LNM scores showed no significant association with language function. For plots displaying all tract-level associations refer to *supplementary figures S7* and *S8*.

The spatial effect patterns, i.e., *β*-coefficient maps, showed significant overlap with 26 of 28 effect pattern pairs being significantly correlated (see *supplementary figure S9* for a correlation matrix).

### Sensitivity analyses

Predictive modelling results were stable when using negative fLNM scores (based on anti-correlations in restingstate fMRI measures) and when including a 25% or 50% thresholding step (*supplementary figure S10*). Exploratory ROI-level inferential statistics based on negative fLNM scores indicated that lower attention / executive function and information processing speed were more significantly associated with more negative fLNM scores in the default mode network (*supplementary figure S11 & S12*).

### Exploratory analyses

Exploratory analyses are detailed in *supplementary text S2*. Functional and structural LNM scores were significantly correlated across ROIs and across subjects (*supplementary figure S13*). Voxel-level lesion network maps indicating white matter regions that contribute to variance in cognitive domain function are shown in *supplementary figure S14 & S15*.

## Discussion

In a large multicentric sample of memory clinic patients, we conducted an in-depth examination of the link between functional and structural LNM scores and cognitive performance. We report two main findings: (1) both structural and functional LNM scores, capturing WMH-related connectivity, significantly improved the prediction of cognitive performance compared to WMH volume; (2) WMH connectivity associated with lower cognitive performance, predominantly mapped to the dorsal and ventral attention networks.

### LNM scores surpass WMH volumes in predicting cognitive performance

In current clinical practice, vascular cognitive impairment in individual patients is often attributed to total WMH burden, but interindividual variance in this relationship can lead to diagnostic dilemmas. Previous research has demonstrated that strategic WMH locations, specifically in commissural and association tracts are statistically more likely to be associated with lower cognitive performance.^4,6,7^ Our approach adds to this perspective, not only considering the localization of WMH but also integrating them with network connectivity information to capture the WMH network embedding. In our analysis, statistical models capitalizing on LNM scores demonstrated superior performance over those relying on total or tract-level WMH volume in predicting cognitive performance in almost all cognitive domains. As this analysis implements current best practices of predictive modelling in neuroimaging, our findings represent evidence for a true prediction of cognitive performance by LNM.^34^ Comparing the improvement from the demographics-based model to the model informed by total WMH volume with the improvement to the model based on both LNM modalities, the usage of fLNM and sLNM scores yielded to a 3-to 7-fold increase in added predictive performance across the three cognitive domains. Moreover, our findings highlighted that total WMH volumes only marginally surpass age, sex, and education in predictive accuracy, stressing the importance of including demographic information as a baseline in predictive models to assess the added value of WMH volume. Collectively, these findings are important, given the longstanding reliance on WMH extent as a primary imaging surrogate marker for cognitive impairment in CSVD. We provide evidence for the considerable role of WMH-related “covert” network effects as indicated previously in studies from smaller clinical or population-based studies.^8,46–48^

Improved prediction of cognitive performance was achieved irrespective of the applied LNM modality. Contrasting prior studies suggesting the inferiority of functional LNM compared to structural approaches for predicting cognitive performance post-stroke,^9,49^ our contrary findings might arise from differences in the LNM approach as well as our focus on WMH rather than ischemic stroke lesions. The ROI-based functional LNM method we used may be more suitable to detect the widespread network disturbances induced by WMH, as opposed to the localized disruptions from stroke lesions. Notably, fLNM and sLNM scores were positively correlated, suggesting some degree of structure-function coupling that could account for their comparable predictive performance. However, the correlation strength was mostly moderate and prediction performance of fLNM and sLNM differed noticeably across sample sizes. In addition, among LNM-informed models, those incorporating both fLNM and sLNM modalities yielded the strongest results. This suggests that both LNM approaches are equally valuable for achieving a high predictive accuracy in general but might also offer complementary information.

Although prediction of almost all cognitive domains was improved by LNM scores, predictive performance for language functions did not exceed that of WMH volumes and demographics. From a network perspective, we argue that this finding can be explained by the relatively confined network of left-lateralized brain regions involved in language functions which might present lower vulnerability to WMH disconnectivity compared to cognitive functions such as information processing speed, that rely on a widely distributed network of brain regions.^50^ In general, the minor improvement of WMH-based measures over the predictive performance attributed to demographics in the whole sample suggests that in this patient population, WMH contribute minimally to the variance in language function.

### WMH related to cognitive impairment map to attention control networks

WMH compromise cognitive performance by impacting the function of specific brain networks. To localize these effects, we investigated *regional* associations between functional and structural LNM scores to cognitive performance. We found that higher LNM scores in cortical areas of the dorsal and ventral attention networks were linked to lower attention and executive function, information processing speed and verbal memory (figure 3). Therefore, higher WMH connectivity in these networks is associated with reduced cognitive performance indicating that WMH impair cognitive function by disrupting the respective connecting white matter fiber tracts.

The dorsal attention network – including the frontal eye field, the superior parietal lobule, the intraparietal sulcus and caudal areas of the medial temporal gyrus – governs top-down attention control by enabling voluntary orientation, with increased activity in response to cues indicating the focus location, timing, or subject.^51,52^ The ventral attention network comprises the frontal and parietal operculum in the inferior frontal gyrus, medial areas of the superior frontal cortex and the temporoparietal junction.^44,53^ This system exhibits activity increases during bottom-up attention control, i.e., upon detection and orientation to salient targets, especially when they appear in unexpected locations.^51,54^ As the effect patterns largely converged on these networks (*supplementary figure S9*), we argue that WMH affect the cognitive functions emerging from these networks, specifically top-down and bottom-up attention control. This aligns with the observation that deficits in attention and executive function are among the most prominent symptoms in CSVD and VCI in general.^1^ Furthermore, prior work demonstrates altered resting-state functional connectivity as well as task activation in attention control networks related to CSVD.^55–57^ Given the covariance of the identified effect patterns, we speculate that WMH contribute to variance in the performance of other cognitive domains, e.g., information processing speed by affecting the attention demands posited by the corresponding tests.

### WMH contribute to cognitive impairment by disrupting frontal and parietal white matter tracts

Regional findings in gray matter areas of the attention control networks are further complemented by white matter tract-level results (figure 4). Functional and structural LNM converged on a significant involvement of tracts connecting frontal and parietal areas involved in attention: the dorsal, medial and ventral section of the superior longitudinal fasciculus – which are known to connect the anterior and posterior parts of the dorsal and ventral attention networks, the medial longitudinal fasciculus, the corticopontine tract, frontoparietal sections of the cingulate, the anterior thalamic radiation, the frontal aslant tract and the arcuate fascicle. Although there were some differences in highlighted tracts between functional and structural LNM, this possibly reflects that both approaches capture different aspects of the same anatomy, with sLNM possibly being more sensitive to direct WMH-induced disruption of axonal connections and functional LNM also reflecting effects mediated via polysynaptic brain circuitry.

Strikingly, in the context of verbal memory, structural WMH connectivity pinpointed a distinct set of memory-relevant tracts: the uncinate fascicle, cingulate, and fornix. Intriguingly, disruptions in fornix connectivity due to WMH were associated with improved verbal memory in patients, a finding that appears counterintuitive given the fornix’s involvement in maintaining memory function. This paradox may be attributable to WMH disrupting inhibitory fibers. For further discussion covering negative fLNM scores/anticorrelations see *supplementary text S16*.

### A unifying hypothesis of WMH disconnectivity

Drawing upon a comprehensive LNM analysis in a memory clinic sample of patients with differing extent and etiology of cognitive impairment, our research converges on a unifying hypothesis: WMH contribute to variance in cognitive functions by disrupting brain circuitry involved in attention control. Our findings not only shed light on the intricate relationships between CSVD, neuroanatomy and cognitive impairment, but they also hint at potential avenues of clinical utilization. The definitive role of CSVD treatments, particularly in precluding cognitive sequelae, is yet to be firmly established. Although there have been promising outcomes related to risk factor modification, particularly blood pressure control,^58,59^ pointing towards enhanced cognitive trajectories, clinical trials in VCI require biomarkers to robustly identify vascular contributions to cognitive impairment and vulnerable individuals. Moving forward, leveraging connectivity information could address this gap contributing to patient-tailored therapeutic interventions and facilitating the identification of subgroups at risk of cognitive disorders through vascular lesions likely to reap the most substantial benefits from medical interventions.

### Strengths and limitations

This study’s strength lies in its integration of innovative analytical techniques with a large, multicentric dataset.^60^ However, we acknowledge several limitations that warrant consideration when interpreting our findings. The inclusion of selected patient samples in several cohorts may limit generalizability to the broader memory clinic population. Additionally, with most patients being of European ancestry, the generalizability of our findings to other ethnicities remains to be established. Furthermore, despite the harmonization of cognitive and imaging data, biases stemming from variations in data acquisition and processing protocols across sites may have impacted our results. On a technical note, while computing fLNM scores, we sampled resting-state BOLD signals in the white matter, typically regarded as noisy and often dismissed as an artifact. However, by integrating it with WMH data, we successfully predicted cognitive performance and demonstrated correlations with structural connectivity information. This challenges the traditional view of the white matter BOLD signal as a mere artefact and supports recent studies – including LNM analyses of white matter lesions in multiple sclerosis – demonstrating that it contains biologically meaningful information.^61–63^

### Conclusion

WMH-related brain network connectivity measures significantly improve the prediction of current cognitive performance in memory clinic patients compared to WMH volume or epidemiological factors. Our findings highlight the contribution of WMH disconnectivity, particularly in attention-related brain regions, to vascular cognitive impairment. As this research field progresses, harnessing neuroimaging markers of WMH connectivity in CSVD has the potential to aid individualized diagnostic and therapeutic strategies.

## Acknowledgements

The authors wish to acknowledge all participants of the Meta VCI Map Consortium. We thank Lei Zhao for his contribution to imaging data harmonization of the Meta VCI Map project data. We thank Guido Cammà and Charlotte M. Verhagen for their contribution to the processing of the imaging data. We thank Olivia K.L. Hamilton, Irene M.C. Huenges Wajer, Bonnie Y.K. Lam, Adrian Wong and Xu Xin as members of the Meta VCI Map neuropsychology working group for their advice on neuropsychological data harmonization.

ADNI data used in preparation of this article were obtained from the Alzheimer’s Disease Neuroimaging Initiative (ADNI) database (adni.loni.usc.edu). As such, the investigators within the ADNI contributed to the design and implementation of ADNI and/or provided data but did not participate in analysis or writing of this report. A complete listing of ADNI investigators can be found at: https://adni.loni.usc.edu/wp-content/uploads/how_to_apply/ADNI_Acknowledgement_List.pdf

