## Supplementary materials for "Enhancing Cognitive Performance Prediction through White Matter Hyperintensity Connectivity Assessment: A Multicenter Lesion Network Mapping Analysis of 3,485 Memory Clinic Patients"

*Supporting Information*

Marvin Petersen, Mirthe Coenen, Charles DeCarli, Alberto de Luca, Ewoud van der Lelij, Alzheimer's Disease Neuroimaging Initiative, Frederik Barkhof, Thomas Benke, Christopher P. L. H. Chen, Peter Dal-Bianco, Anna Dewenter, Marco Duering, Christian Enzinger, Michael Ewers, Lieza G. Exalto, Evan F. Fletcher, Nicolai Franzmeier, Saima Hilal, Edith Hofer, Huberdina L. Koek, Andrea B. Maier, Pauline M. Maillard, Cheryl R. McCreary, Janne M. Papma, Yolande A. L. Pijnenburg, Reinhold Schmidt, Eric E. Smith, Rebecca M. E. Steketee, Esther van den Berg, Wiesje M. van der Flier, Vikram Venkatraghavan, Narayanaswamy Venketasubramanian, Meike W. Vernooij, Frank J. Wolters, Xu Xin, Andreas Horn, Kaustubh R. Patil, Simon B. Eickhoff, Götz Thomalla, J. Matthijs Biesbroek, Geert Jan Biessels, Bastian Cheng

### Methods

#### Supplementary figure S1 – Investigated white matter tracts of the HCP1065 atlas

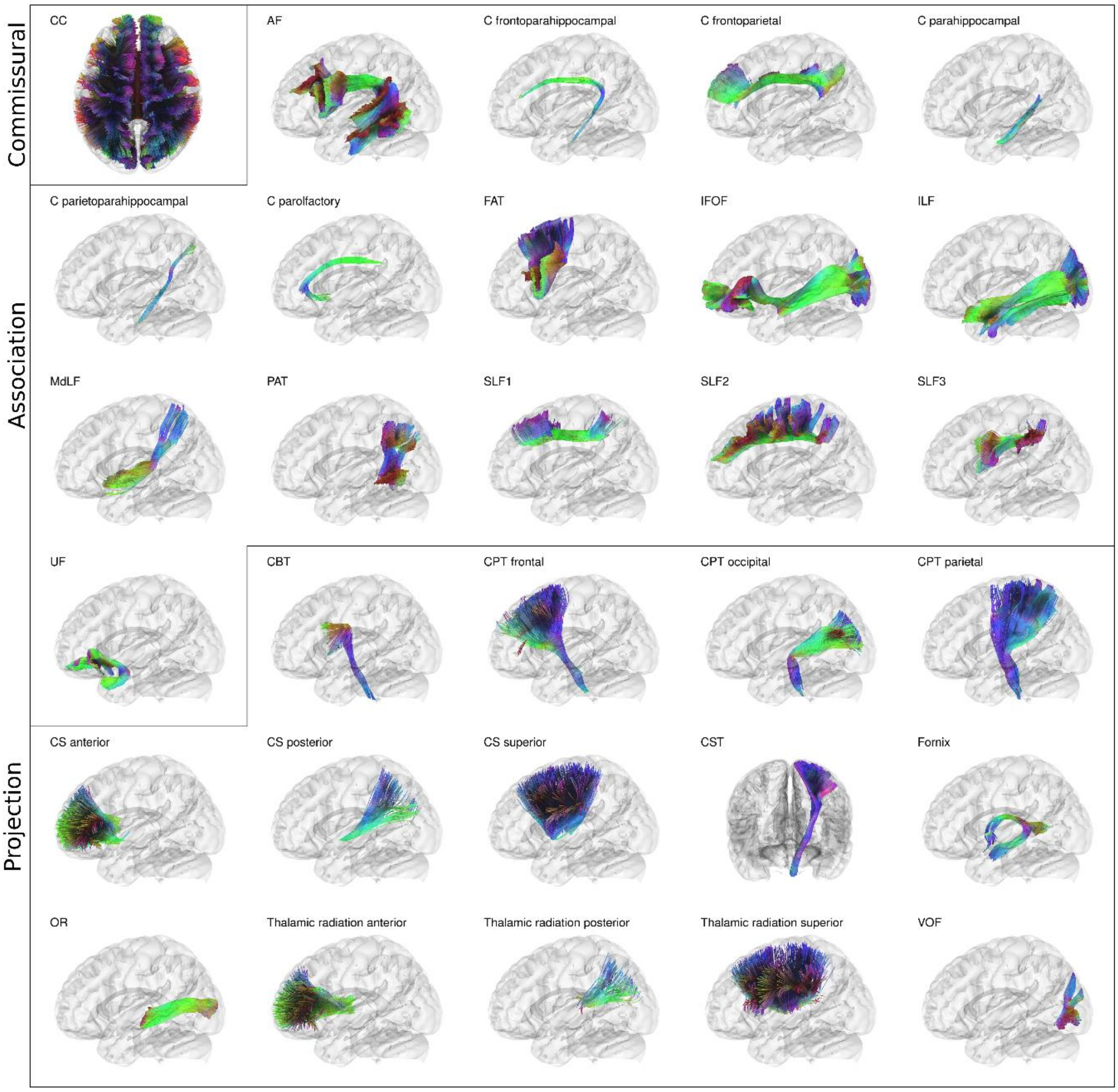

Anatomical depiction of the white matter tracts investigated, categorized into association, projection and commissural tracts. For paired tracts only left side examples are visualized. *Tract abbreviations*: Commissural tracts – CC = corpus callosum; Association tracts - AF = arcuate fascicle, C = cingulate, FAT = frontal aslant tract, IFOF = inferior fronto-occipital fasciculus, ILF = inferior longitudinal fasciculus, MdLF = middle longitudinal fasciculus, PAT = posterior aslant tract, SLF = superior longitudinal fasciculus, UF = uncinate fasciculus; Projection tracts – CBT = corticobulbar tract, CPT = corticopontine tract, CS = corticostriatal pathway, CST = corticospinal tract, FPT = frontopontine tract, F = fornix, OPT = occipitopontine tract, OR = optic radiation, VOF = Vertical occipital fasciculus.

#### Supplementary text S2 - Exploratory analyses

##### Correlation of lesion network mapping scores

To test for a structure-function-coupling of lesion network mapping scores, we correlated functional and structural lesion network mapping scores 1) across subjects per region of interest and 2) across regions of interests per subjects. Corresponding results can be found in *supplementary figure S13.*

##### Voxel-level lesion network maps

In addition, we generated voxel-level lesion network maps to identify white matter areas crucial for cognitive performance. This involved averaging the voxel-level connectivity maps of the ROIs significantly linked to cognitive domain scores. These maps, created for each combination of the four cognitive domains and two LNM modalities, highlight regions where connectivity links to cognitive variance. We then scaled these maps by the WMH frequency map, which reflects the prevalence of WMH in each voxel across the analysis sample. The resulting maps reveal regions where WMH most commonly contribute to variance in cognitive performance. The maps can be found in *supplementary figures S14 & S15.*

### Results

#### Figure S3 – White matter hyperintensity distribution

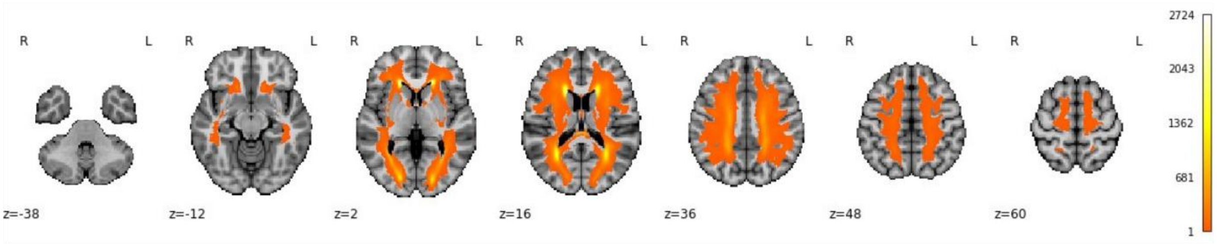

Heatmap indicating the frequency of white matter hyperintensities across the analysis sample.

#### Figure S4 – Predictive modelling analysis with explained variance ($R^2$, coefficient of determination) scoring

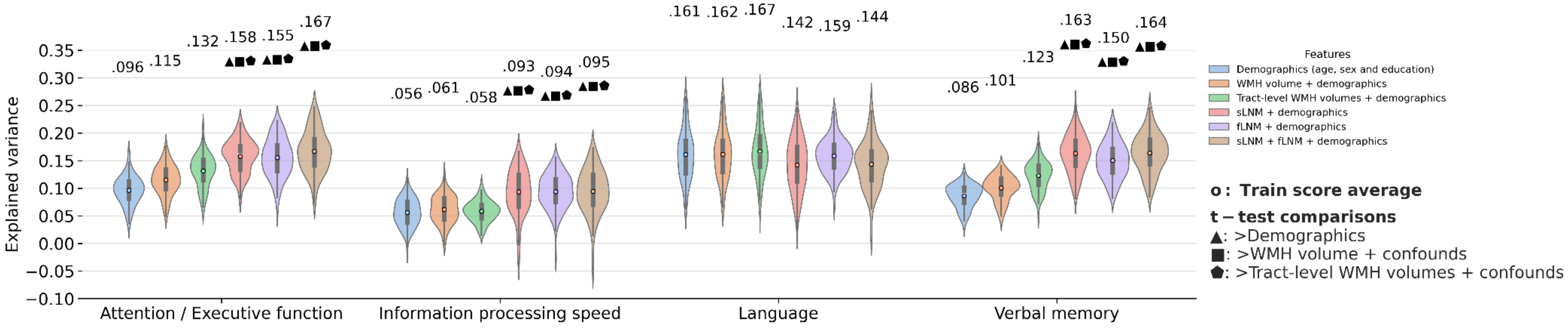

Violin plots illustrate prediction outcomes across cognitive domains. Each violin displays the distribution of explained variance of cognitive domain scores (10-fold cross-validation x 10 repeats = 100 folds → 100 Pearson correlations) for a model informed by a different feature set. The higher the explained variance, the higher the prediction performance. Blue: confounds (age, sex and education); orange: total WMH volume + confounds; green: tract-level WMH volumes + confounds; red: sLNM scores + confounds; purple: fLNM scores + confounds; brown: sLNM scores + fLNM scores + confounds. The average explained variance is indicated above each violin, with colored dots showing training score averages. Geometric symbols denote t-test results comparing LNM-based models against confound- and WMH volume-based models: ▲ indicates higher explained variance than confounds, ■ than WMH volume + confounds, ⬟ than tract-level WMH volume + confounds. Of note, a negative explained variance is possible using sum-of-squares formulation. A negative value indicates that the optimized model fits the data worse than a horizontal line representing the mean of the target variable.

#### Table S5 – Predictive modelling analysis results – Average negative mean squared error

| | Attention / executive function | Information processing speed | Language | Verbal memory |
|---|---|---|---|---|
| Confounds (age, sex education) | -1.06219 | -2.43104 | -2.91271 | -1.51663 |
| WMH volume + confounds | -1.03992 | -2.41763 | -2.9114 | -1.49302 |
| Tract-level WMH volumes + confounds | -1.02051 | -2.42529 | -2.8931 | -1.45587 |
| sLNM + confounds | -0.98846 | -2.33568 | -2.98366 | -1.38662 |
| fLNM + confounds | -0.99139 | -2.33465 | -2.92767 | -1.40741 |
| sLNM + fLNM + confounds | -0.97774 | -2.33223 | -2.97935 | -1.38486 |

### Figure S6 – Region of interest-level averages of lesion network mapping scores

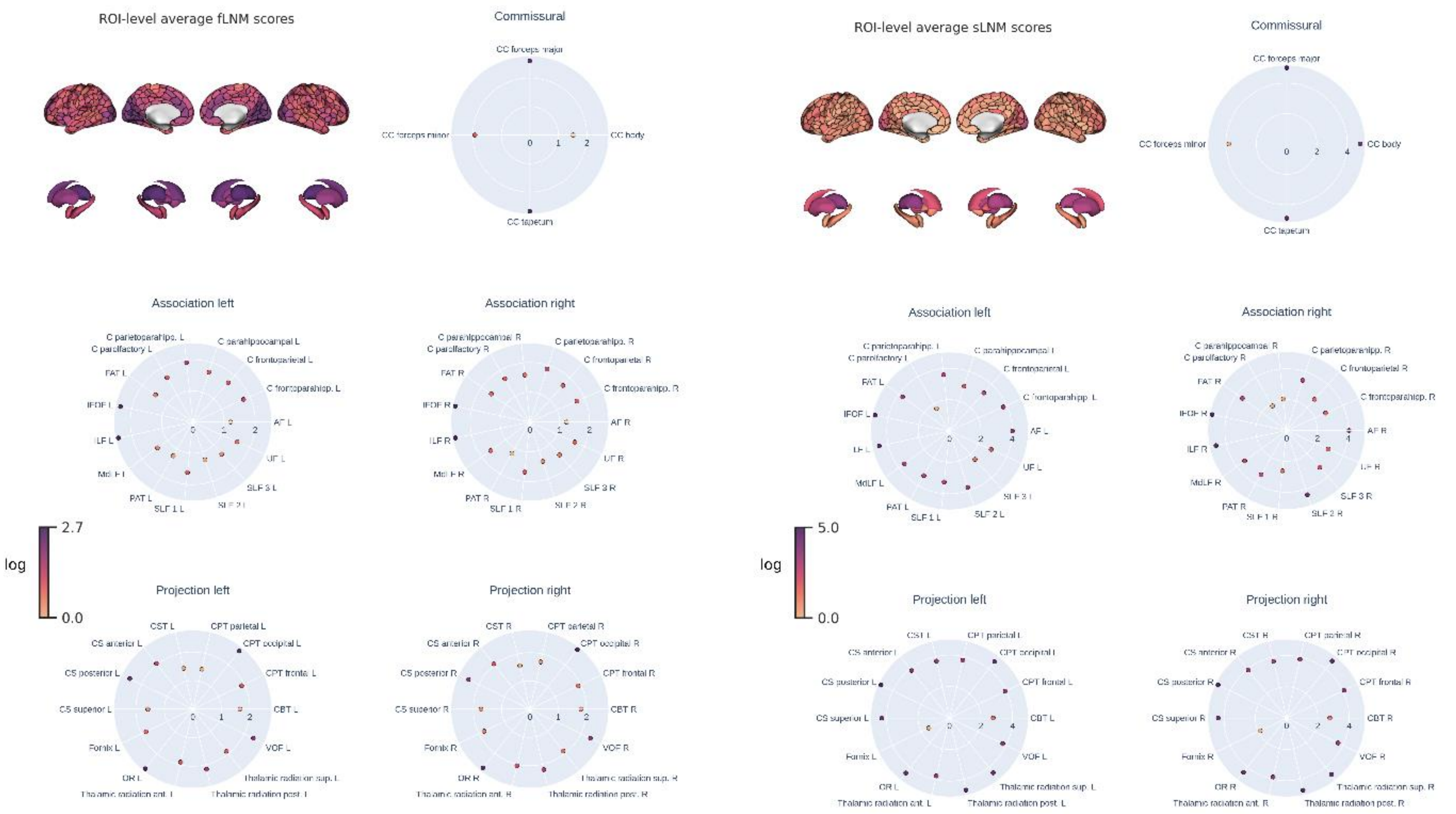

Region of interest-level average functional and structural lesion network mapping scores. Average lesion network mapping scores of gray matter regions of interest are mapped on the cortical surface and subcortical regions. For the white matter tracts, average lesion network mapping scores are displayed in radar plots. Radar plots display white matter tracts in alphabetical order starting at the 3 o'clock position.

#### Figure S7 – Tract-level functional lesion network mapping

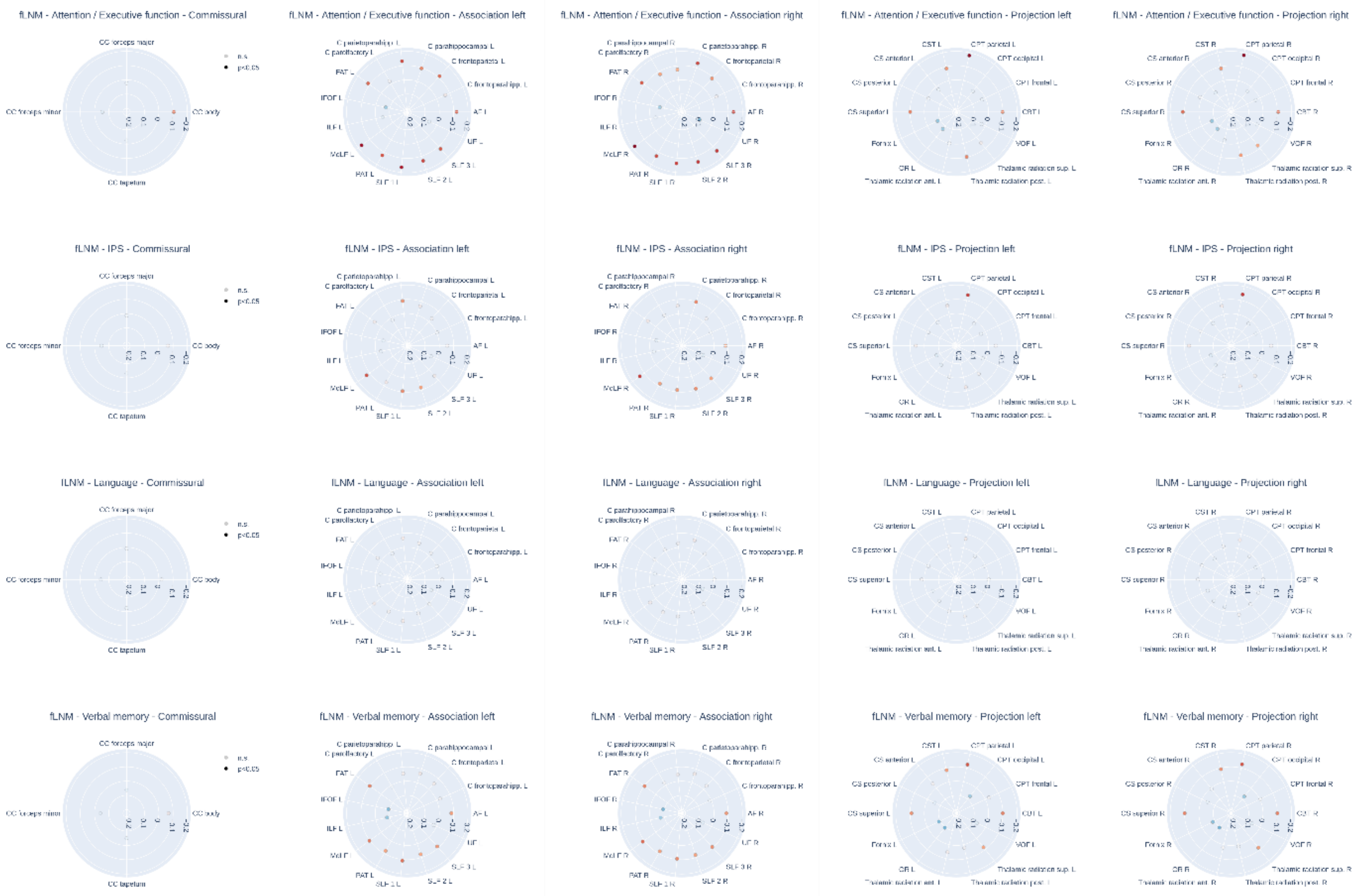

Radar plots display tract-level β coefficients from inferential statistics indicating the relationship between regional functional lesion network mapping scores and cognitive domain scores. This plot shows the associations for all tracts while in the main manuscript only the top 10 effects per combination of LNM modality and cognitive domain are featured. In contrast to the main manuscript, tracts are displayed in alphabetical order starting at the 3 o'clock position in the counterclockwise position. Red dots indicate a negative association (higher LNM score – lower cognitive domain score) and blue dots indicate a positive association (higher LNM score – higher cognitive domain score). Faintly colored dots indicate non-significant associations. Tracts with a significant association are displayed below the radar plots in alphabetical order. For paired tracts only left side examples are visualized.

### Figure S8 – Tract-level structural lesion network mapping

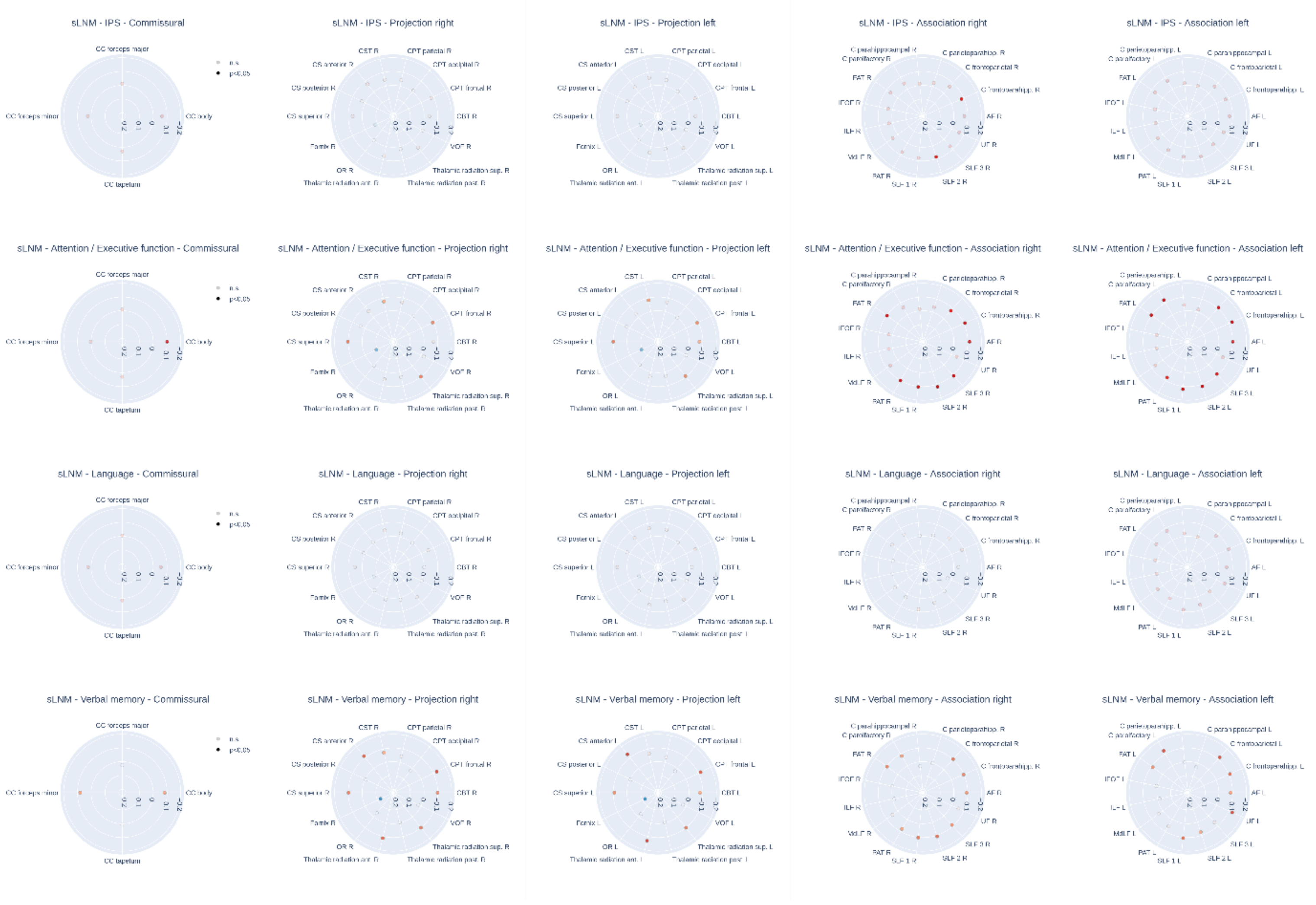

Radial plots display tract-level β coefficients from inferential statistics indicating the relationship between regional structural lesion network mapping scores and cognitive domain scores. This plot shows the associations for all tracts while in the main manuscript only the top 10 effects per combination of LNM modality and cognitive domain are featured. In contrast to the main manuscript, tracts are displayed in alphabetical order starting at the 3 o'clock position in the counterclockwise position. Red dots indicate a negative association (higher LNM score – lower cognitive domain score) and blue dots indicate a positive association (higher LNM score – higher cognitive domain score). Faintly colored dots indicate non-significant associations. Tracts with a significant association are displayed below the radar plots in alphabetical order. For paired tracts only left side examples are visualized.

#### Figure S9 – Spatial correlations of region of interest-level β coefficients

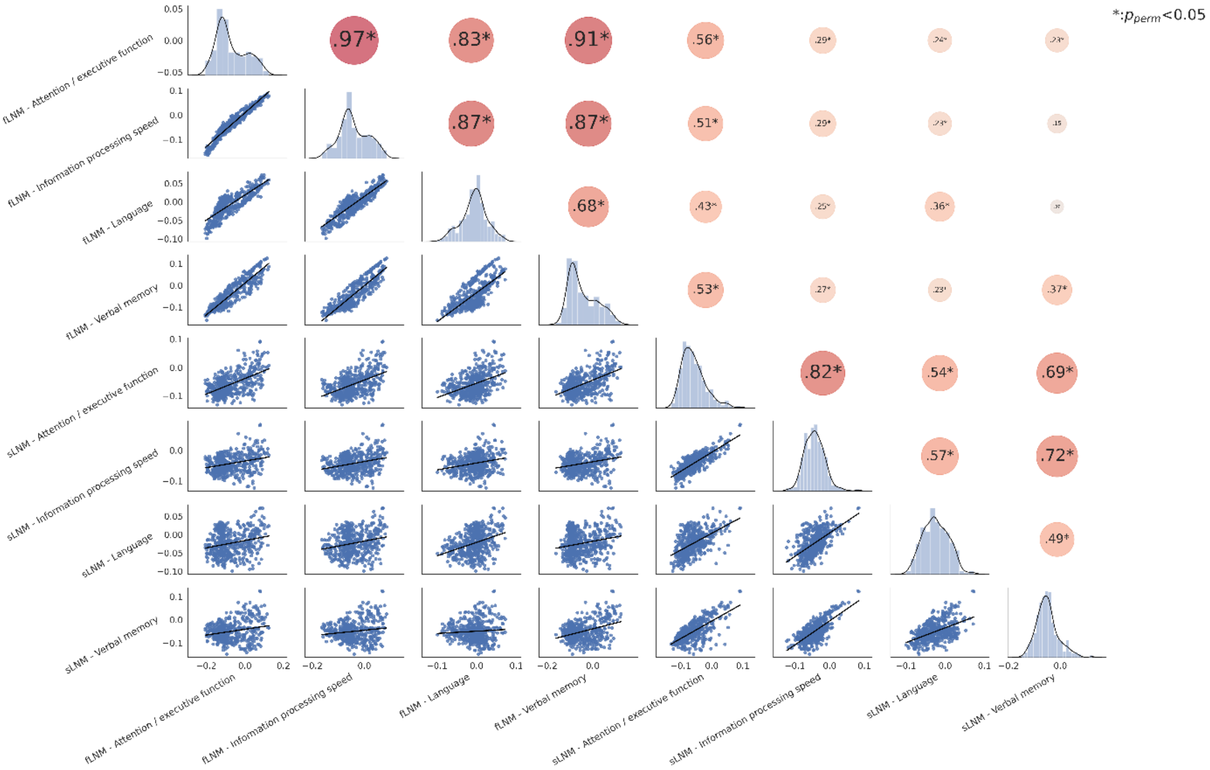

Spatial correlation matrix of all ROI-level effect maps ($\beta$). To investigate the spatial correspondence between effect maps of the ROI-level analysis, we performed Spearman correlations of each pair of maps. The upper triangle of the matrix displays spearman correlations with dot size and color representing the orientation and magnitude of the correlation coefficients. Asterisks highlight significant correlations after permutation testing and false discovery rate correction. The diagonal shows kernel density plots. The lower triangle illustrates the linear relationships via regression plots. Each dot of the regression plot corresponds with a ROI. *Abbreviations*: fLNM = functional lesion network mapping, $p_{perm}$ = p-value obtained via comparison of empirical Spearman correlation to permutation-based null distribution, ROI = region of interest, $r_{sp}$ = Spearman correlation, sLNM = structural lesion network mapping.

#### Figure S10 – Sensitivity analysis: Predictive modelling analysis

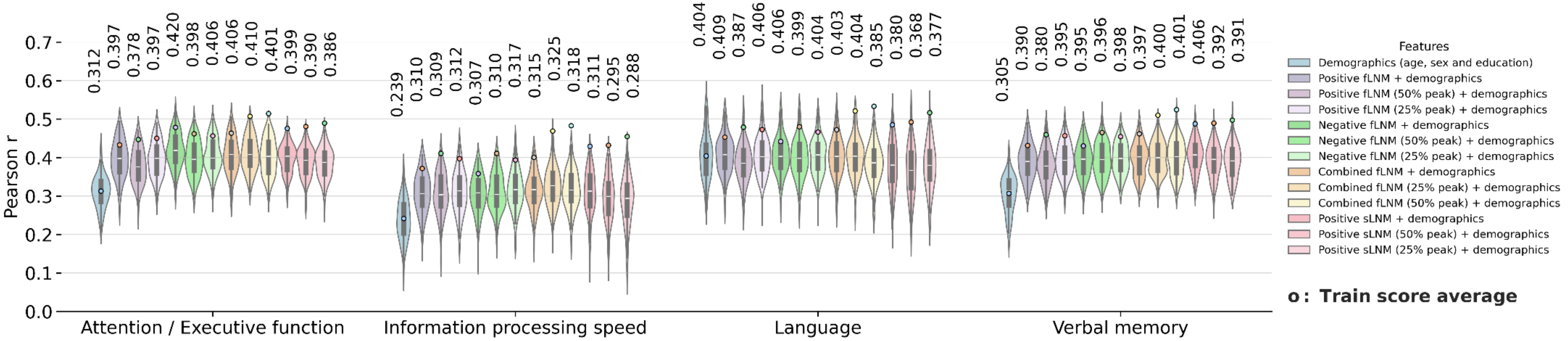

This plot corresponds with Figure 2 of the main manuscript but displays model performances informed by negative fLNM scores as well as LNM scores computed via different thresholding schemes alongside original LNM-informed models. Negative fLNM scores were obtained by only considering negative Pearson correlation coefficients within the WMH mask. Thresholding was performed by averaging only the highest 25% (25% peak) and highest 50% (50% peak) of intensity values of the ROI-level connectivity map in the WMH mask. For the negative fLNM scores, the lowest 25% and 50% voxel intensity values were averaged instead. Combined fLNM indicates models informed by positive and negative fLNM scores. *Abbreviations*: fLNM = functional lesion network mapping, sLNM = structural lesion network mapping, WMH = white matter hyperintensities of presumed vascular origin.

#### Figure S11 – Sensitivity analysis: Inferential statistics results of cortical and subcortical gray matter based on negative functional lesion network mapping scores

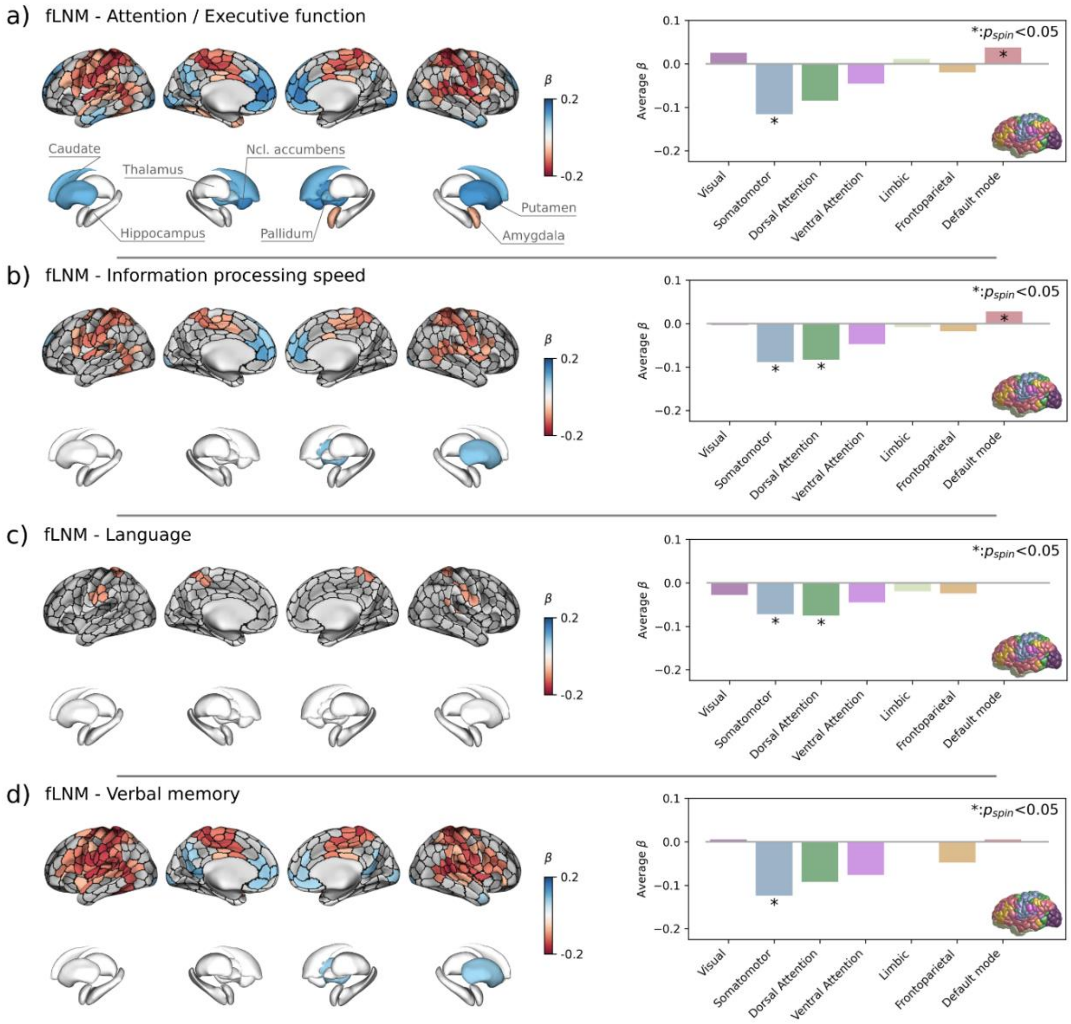

This plot corresponds with Figure 3 a) – d) of the main manuscript but in contrast displays regional associations of fLNM scores based on anticorrelations. Left: ROIs that were significantly associated with cognitive domain scores after family wise error-correction are highlighted by colors encoding $\beta$-coefficients from general linear models: a negative $\beta$ (red) denotes that a higher regional LNM score, i.e., higher WMH connectivity, is associated to a lower cognitive domain performance; a positive $\beta$ (blue) indicates that a higher regional LNM score is linked to a higher cognitive domain performance. Right: Barplots displaying the average $\beta$ in the canonical (Yeo) resting state networks. The brain on the right indicates the regional distribution of the canonical resting state networks with colors corresponding to the bars. Statistical significance was assessed using spin permutations. Each row corresponds with a different combination of lesion network mapping modality and cognitive domain: a) fLNM – attention / executive function, b) fLNM – information processing speed, c) fLNM – language, d) fLNM – verbal memory. *Abbreviations*: fLNM = functional lesion network mapping, $p_{spin}$ = p-value derived from spin permutations, ROIs = regions of interest, sLNM = structural lesion network mapping.

#### Figure S12 – Sensitivity analysis: Inferential statistics results of white matter tracts based on negative functional lesion network mapping scores

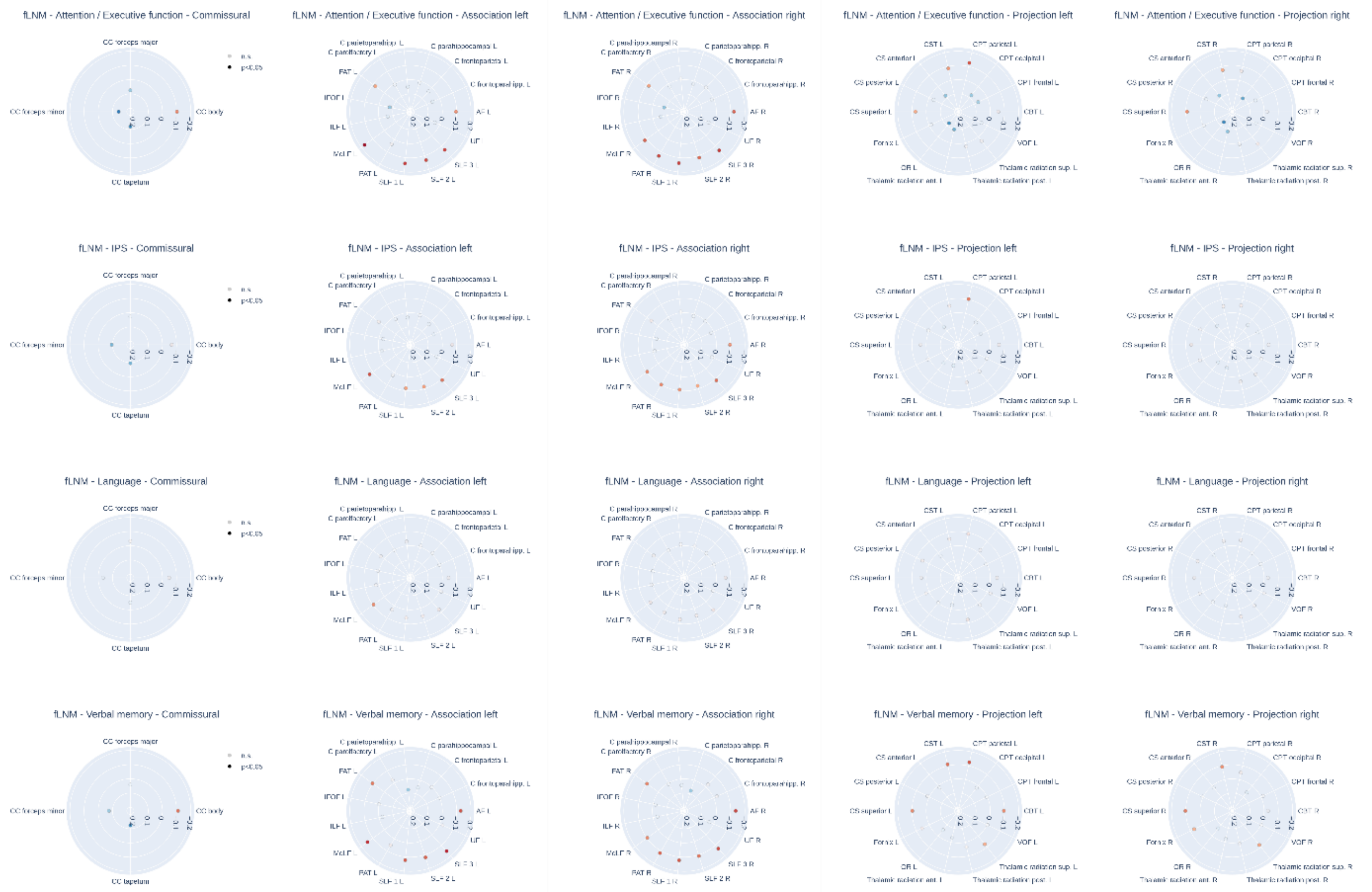

This plot corresponds with Figure 4 of the main manuscript but in contrast displays regional associations of fLNM scores based on anticorrelations. Tract abbreviations: Commissural tracts – CC = corpus callosum; Association tracts - AF = arcuate fascicle, C = cingulate, FAT = frontal aslant tract, IFOF = inferior fronto-occipital fasciculus, ILF = inferior longitudinal fasciculus, MdLF = middle longitudinal fasciculus, PAT = posterior aslant tract, SLF = superior longitudinal fasciculus, UF = uncinate fasciculus; Projection tracts – CBT = corticobulbar tract, CPT = corticopontine tract, CS = corticostriatal pathway, CST = corticospinal tract, CT = corticothalamic pathway, FPT = frontopontine tract, F = fornix, OPT = occipitopontine tract, OR = optic radiation, VOF = Vertical occipital fasciculus. *Abbreviations*: fLNM = functional lesion network mapping, n.s. = non-significant, p = p-value, sLNM = structural lesion network mapping.

#### Figure S13 – Structure-function correlations of regional lesion network mapping scores

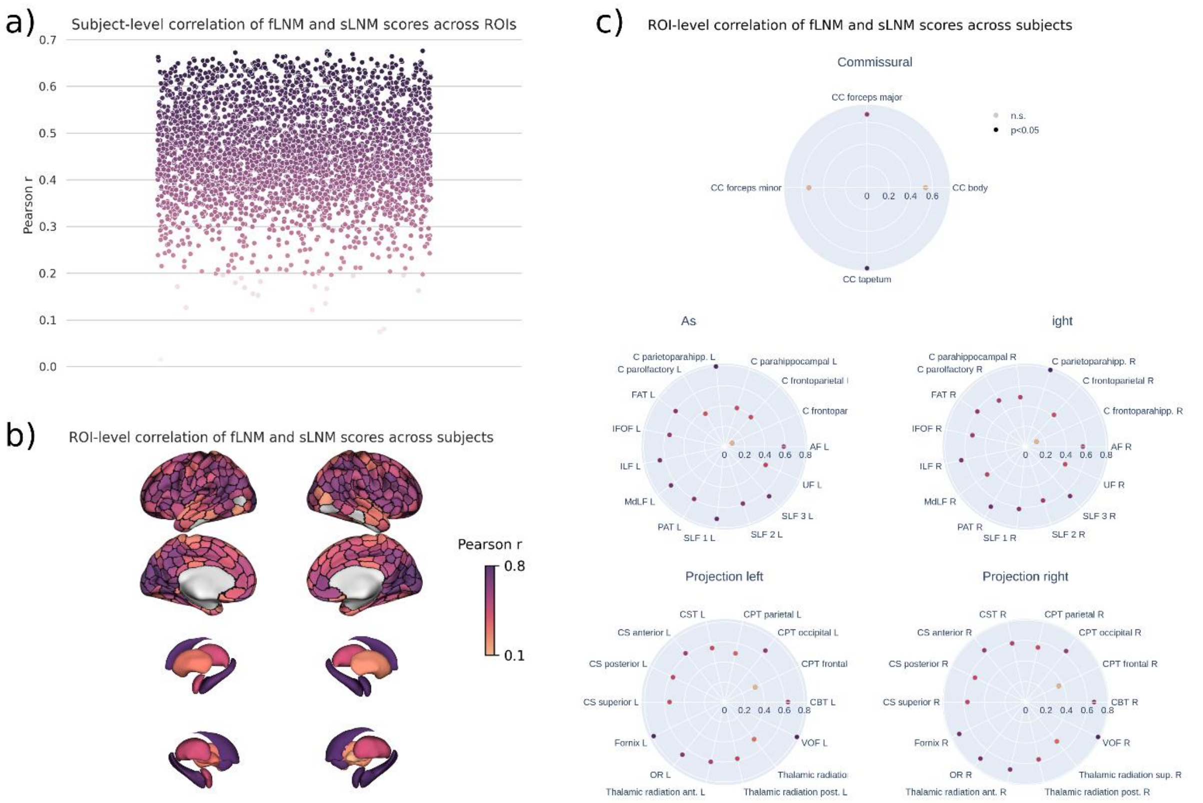

a) Swarmplot displaying the Pearson correlation of fLNM and sLNM scores across ROIs per subject. Each dot represents a subject and is colored by the Pearson correlation. b) and c) Pearson correlation of fLNM and sLNM scores across subjects per ROI. *Abbreviations*: fLNM = functional lesion network mapping, ROI = region of interest, sLNM = structural lesion network mapping.

#### Figure S14 - Voxel-level lesion network maps

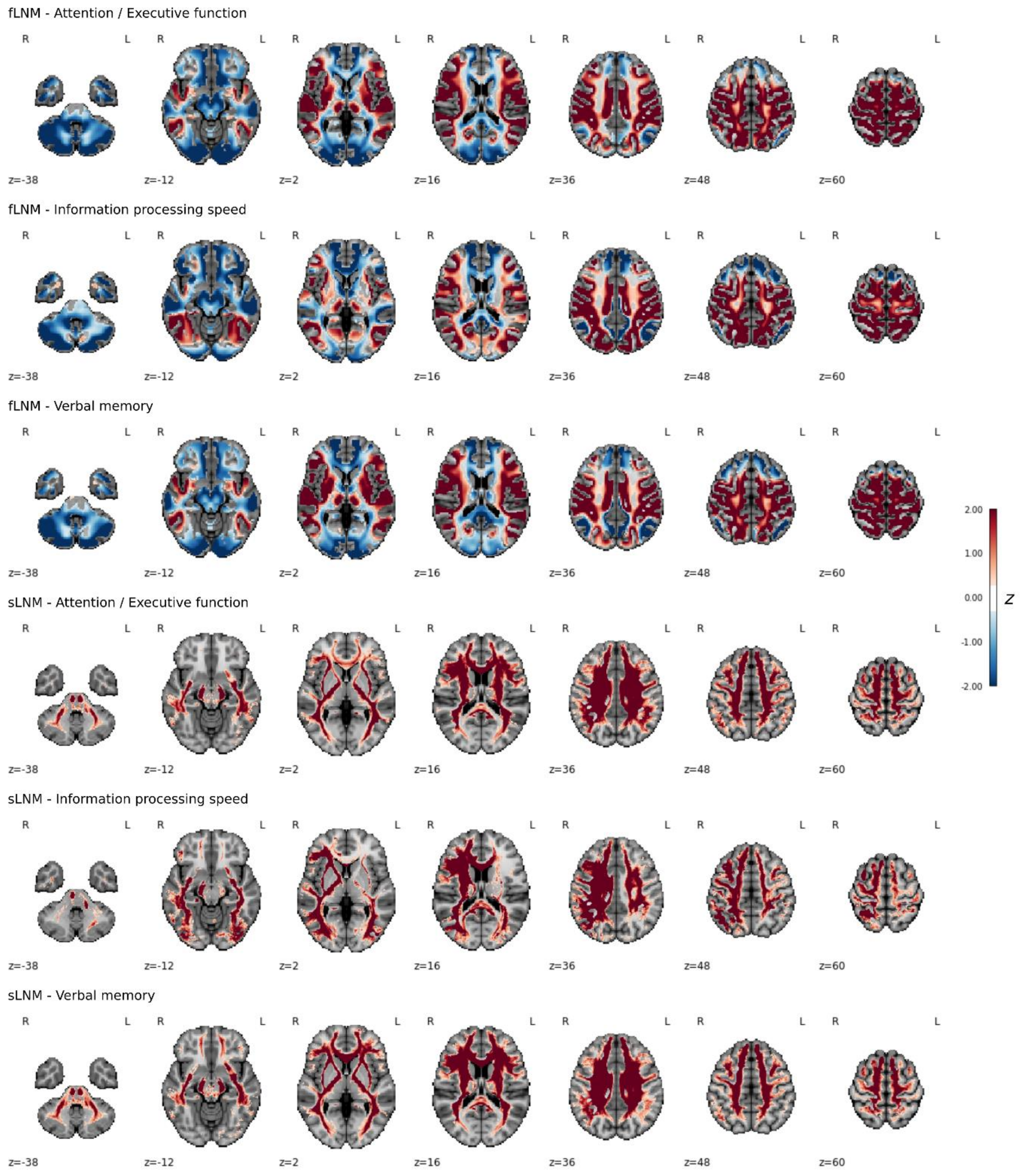

Voxel-level lesion network maps indicate the connectivity to regions of interest that significantly contribute to a cognitive domain. Each row corresponds to a different combination of lesion network mapping modalities (functional and structural) and cognitive domain scores. For the functional lesion network maps, positive z-scores indicate a positive Pearson correlation with the resting-state BOLD signal of the significantly associated ROIs. Negative z-scores indicate anticorrelated voxels and are highlighted in blue. For the structural lesion network maps, deeper red indicates that a voxel is connected by a higher amount of streamlines to significantly associated ROIs. *Abbreviations*: ROI = region of interest.

#### Figure S15 - Voxel-level lesion network maps scaled by the white matter hyperintensity distribution map

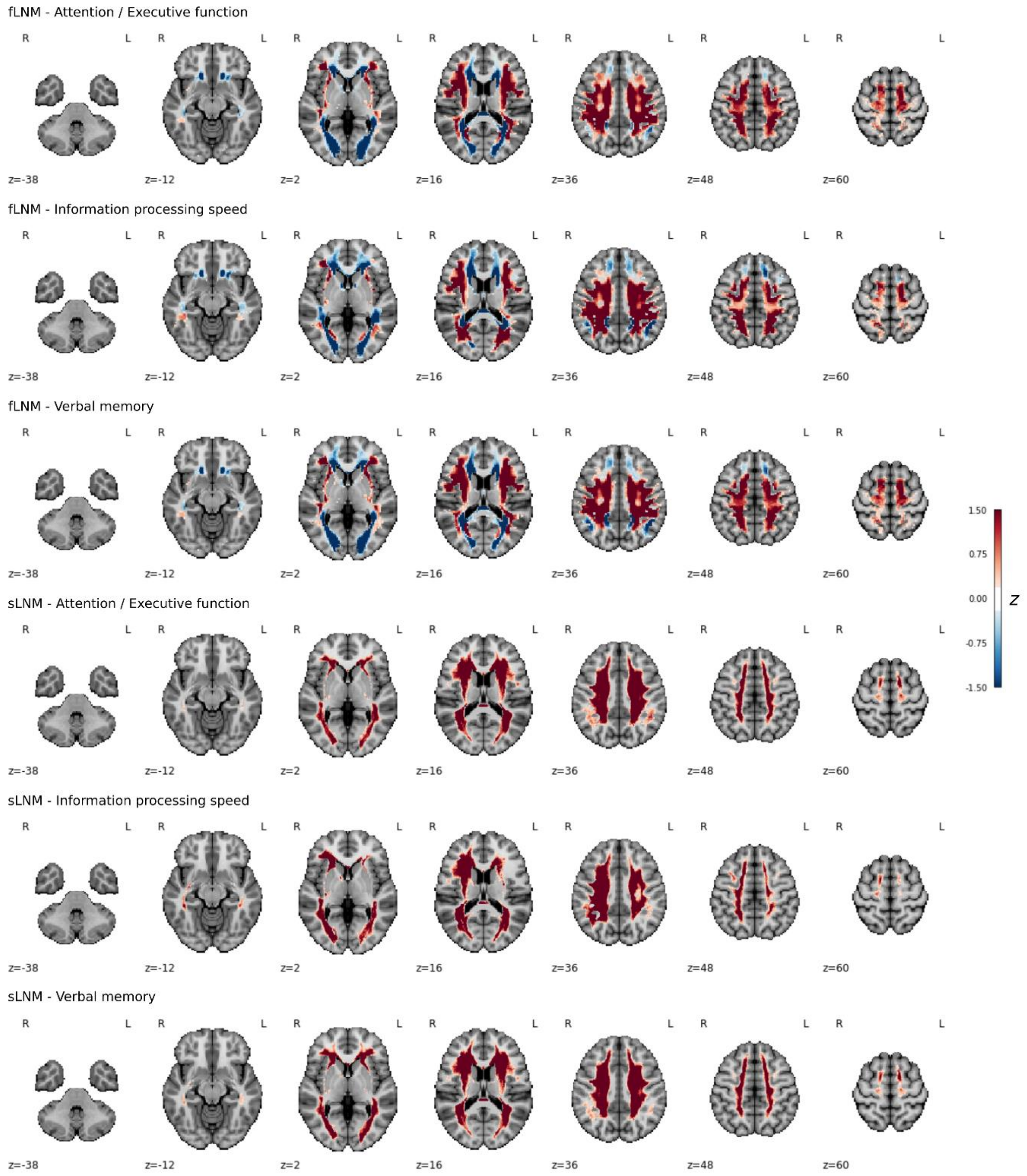

Voxel-level lesion network maps scaled by the WMH frequency map indicate the connectivity to regions of interest that significantly contribute to a cognitive domain and are likely lesioned by WMH. *Abbreviations*: ROI = region of interest, WMH = white matter hyperintensities of presumed vascular origin.

#### Discussion

##### Text S16 – Negative functional lesion network mapping scores / anticorrelations

The attention control networks are functionally contrasted by the default mode network which shows, instead of being engaged during externally focused tasks, increased activity during internally directed attention and self-referential processes.[1] As a result, the default mode network and the attention control networks are often found to be anticorrelated at rest.[2] This anticorrelation is thought to reflect a fundamental aspect of brain organization and the complex dynamic interplay between the networks is thought to be central for cognitive processing. Resting-state fMRI studies in CSVD patients suggest that WMH might affect the DMN and attention network interaction, particularly affecting anterior-posterior communication by disrupting long associative white matter fiber tracts.[3,4] Our findings indicate that stronger anticorrelation between the default mode network and WMH – reflected by more negative fLNM scores – correlates with reduced attention, executive function, and processing speed, supporting this hypothesis. Furthermore, by demonstrating enhanced predictive performance based on negative fLNM scores our results underscore the perception of anticorrelations yielding biologically and clinically meaningful information.
